# Connectivity of the Piriform Cortex and its Implications in Temporal Lobe Epilepsy

**DOI:** 10.1101/2024.07.21.24310778

**Authors:** Alfredo Lucas, Marc Jaskir, Nishant Sinha, Akash Pattnaik, Sofia Mouchtaris, Mariam Josyula, Nina Petillo, Rebecca W. Roth, Gulce N. Dikecligil, Leonardo Bonilha, Jay Gottfried, Ezequiel Gleichgerrcht, Sandhitsu Das, Joel M. Stein, James J. Gugger, Kathryn A. Davis

**Affiliations:** Perelman School of Medicine, University of Pennsylvania; Department of Bioengineering, University of Pennsylvania; Neuroscience Graduate Group, University of Pennsylvania; Department of Neurology, University of Pennsylvania; Department of Neurology, University of South Carolina; Department of Radiology, University of Pennsylvania; Department of Neurology, Emory University

## Abstract

**Background:** The piriform cortex has been implicated in the initiation, spread and termination of epileptic seizures. This understanding has extended to surgical management of epilepsy, where it has been shown that resection or ablation of the piriform cortex can result in better outcomes. How and why the piriform cortex may play such a crucial role in seizure networks is not well understood. To answer these questions, we investigated the functional and structural connectivity of the piriform cortex in both healthy controls and temporal lobe epilepsy (TLE) patients.

**Methods:** We studied a retrospective cohort of 55 drug-resistant unilateral TLE patients and 26 healthy controls who received structural and functional neuroimaging. Using seed-to-voxel connectivity we compared the normative whole-brain connectivity of the piriform to that of the hippocampus, a region commonly involved in epilepsy, to understand the differential contribution of the piriform to the epileptogenic network. We subsequently measured the inter-piriform coupling (IPC) to quantify similarities in the inter-hemispheric cortical functional connectivity profile between the two piriform cortices. We related differences in IPC in TLE back to aberrations in normative piriform connectivity, whole brain functional properties, and structural connectivity.

**Results:** We find that relative to the hippocampus, the piriform is functionally connected to the anterior insula and the rest of the salience ventral attention network (SAN). We also find that low IPC is a sensitive metric of poor surgical outcome (sensitivity: 85.71%, 95% CI: [19.12%, 99.64%]); and differences in IPC within TLE were related to disconnectivity and hyperconnectivity to the anterior insula and the SAN. More globally, we find that low IPC is associated with whole-brain functional and structural segregation, marked by decreased functional small-worldness and fractional anisotropy.

**Conclusions:** Our study presents novel insights into the functional and structural neural network alterations associated with this structure, laying the foundation for future work to carefully consider its connectivity during the presurgical management of epilepsy.

## Introduction

In epilepsy, particularly temporal lobe epilepsy (TLE), the piriform cortex has emerged as a significant focus of study. TLE is the most prevalent form of epilepsy in adults, characterized by seizures that are often drug-resistant^1–3^. The piriform cortex, which has a biological analogue in rodents and non-human primates termed the “area tempestas”, has been implicated in the initiation, spread and termination of epileptic seizures, acting as a critical hub in the epileptogenic network^4–8^. This is not surprising as the piriform is phylogenetically similar to the hippocampus, a key node in epileptogenic networks, and also has broad connections to limbic and cortical structures^9^. This understanding has extended to surgical management of epilepsy, where it has been shown that resection or ablation of the piriform cortex can result in better outcomes^10–15^.

How and why the piriform cortex may play such a crucial role in seizure networks is not well understood. Studies have demonstrated functional connectivity abnormalities that localize to the piriform cortex, even when seizures arise from remote brain areas^16–19^, however, a detailed understanding of the structural and functional connectivity of the human piriform cortex to the rest of the brain is lacking. Furthermore, how aberrations in the structural and functional connectivity of the piriform cortex relate to TLE and surgical outcomes is not known. This stands in stark contrast to the hippocampus, whose functional and structural connectivity with the rest of the brain, as well as its association with the underlying epileptogenic network, have been extensively studied^20–27^. Therefore, we aimed to explore the relationship between the piriform cortex and the epileptogenic network in TLE by examining its functional and structural connectivity with the rest of the brain.

In this study, we hypothesized that the piriform cortex exhibits a distinct whole-brain connectivity profile compared to the hippocampus, and that its connectivity to normative brain regions is impaired in temporal lobe epilepsy (TLE). Given that olfaction, the primary sensory function of the piriform cortex, is a highly bilateral sense^28–30^, we further hypothesized that cortical connectivity patterns between piriform cortices would be discordant in TLE. We anticipate that differences in inter-piriform coupling in TLE are driven by compromised connectivity to the normative piriform network. Such differences are expected to reflect broader whole-brain functional and structural connectivity patterns, indicative of differential integration and segregation within the entire brain network in TLE.

To answer these questions, we investigated the functional and structural connectivity of the piriform cortex using seed-to-voxel functional connectivity and diffusion tractography in both healthy controls and TLE patients. We compared the connectivity patterns of the piriform cortex to the hippocampus to understand the unique contribution of the piriform to seizure networks. Next, we measured the inter-piriform coupling (IPC) to quantify similarities in the inter-hemispheric cortical functional connectivity profile between the two piriform cortices. We find that (a) relative to the hippocampus, the piriform is functionally connected to a separate canonical brain network. We also find that (b) the IPC is associated with positive surgical outcomes; and differences in IPC within TLE are related to functional properties within the normative piriform network identified in controls, as well as structural connectivity abnormalities. Our results provide a detailed systematic analysis of piriform cortex connectivity, how the connectivity is altered in relationship to TLE, and its association with epilepsy surgical outcomes.

## Methods

### Participant Demographics

Data acquisition for this study was approved by the institutional review board of the University of Pennsylvania. We used a retrospective cohort of 55 drug-resistant unilateral TLE patients who underwent neuroimaging during presurgical evaluation at the Hospital of the University of Pennsylvania.

The Penn Epilepsy Surgical Conference (PESC) determined the location of seizure focus for all patients using a multidisciplinary approach, incorporating clinical, neuroimaging, and neurophysiological factors such as seizure semiology, brain MRI, FDG-PET, MEG, scalp EEG, and intracranial EEG. The PESC team consisted of neurologists, neurosurgeons, neuropsychologists, neuroradiologists, and nuclear medicine specialists. Thirty-four patients had left-sided lateralization and 21 had right-sided lateralization of the seizure focus. The study also included 26 healthy control participants. Demographic information, MRI lesional status, final lateralization, and Engel surgical seizure outcomes at 24 months, where available, are recorded in **Table 1**. We additionally included a subset of 100 unrelated Human Connectome Project (HCP) participants, and epilepsy (n=29) and control (n=10) subjects from an Emory university dataset as validation (**Supplementary Table 2**).

**Table 1.** Subject Demographics.

Subject Demographics
| Characteristic | Healthy Controls | TLE |
| --- | --- | --- |
| **Total Subjects** | 26 | 55 |
| **Age** | 30±10 | 39±11 |
| **Female** | 12 | 31 |
| **Disease Duration (years)** | - | 17±15 |
| **Lesional MRI** | - | 35 |
| **Disease Laterality** |  |  |
| *Left* | - | 34 |
| *Right* | - | 21 |
| **Surgical Outcomes (24 months)** |  |  |
| *Engel IA* | - | 7 |
| *Engel IB-IV* | - | 20 |
| *Unavailable* | - | 36 |
| **Acquisition Protocol** |  |  |
| *Version A* | 7 | 10 |
| *Version B* | 19 | 45 |

### Functional and Structural MRI Acquisition

Two different fMRI acquisition protocols were followed (fMRI protocols A and B; see harmonization details in the *Seed-to-voxel Functional Connectivity* section). The number of subjects that were scanned with each protocol is shown in Table 1. fMRI images for protocol A were acquired with 3.0 mm isotropic voxel size, echo time (TE)/repetition time (TR) = 30/500 ms, a multiband factor of 6 and a 7-min acquisition time. fMRI images for protocol B were acquired with a 2.0 mm isotropic voxel size, TE/TR = 37/800 ms, a multiband factor of 6 and a 9-min acquisition time. For both protocols, T1 weighted images were acquired with a sagittal, 208-slice MPRAGE sequence, TE/TR = 2.24/2400 ms, inversion time (TI) =1060 ms, field-of-view (FOV) = 256mm, with a 0.8 isotropic voxel size.

### Diffusion Weighted MRI acquisition

Diffusion-weighted MRI data were acquired on a 3T Siemens Prisma scanner using a single-shot echo planar imaging multi-shell sequence (116 diffusion sampling directions, b-values of 0, 300, 700, and 2000 s/mm2, resolution = 2 x 2 x 2 mm3, field of view = 220 mm, TR = 4.3 ms, TE = 75 ms). To correct for B0 inhomogeneities in DWI acquisition, we acquired top-up sequences with reverse phase encoding directions or phase and magnitude images.

### Functional Image Preprocessing

BOLD rs-fMRI from the Penn and Emory datasets were preprocessed following the same pipeline as Lucas et. al.^31^ Briefly, using a combination of fMRIprep^32^ and XCP-Engine^33^, we demeaned, detrended, regressed confounds using a 36-parameter model (translation and rotation in 3 directions, white matter, cerebrospinal fluid, global signal, their first derivative and their square), bandpassed, and co-registered to the 152 non-linear asymmetric 2009 Montreal Neurological Institute (MNI) version c template space each subject’s rs-fMRI acquisition. The PIER dataset was corrected for susceptibility distortion using acquired fieldmaps, whereas the Emory dataset was corrected using a fieldmap-less approach provided with fMRIprep.

We also included the ICA-FIX (hp2000_clean) preprocessed Human Connectome Project (HCP) rs-fMRI data from 100 unrelated subjects^34^. Since these data are already motion regressed, we added the white matter, cerebrospinal fluid, global signal, their first derivative and their square as additional confounds.

### Diffusion Weighted MRI Preprocessing

We ran the FreeSurfer “recon-all” pipeline (version 7.1) on preoperative T1-weighted MRI to generate gray and white-matter surfaces. Freesurfer generated surfaces were quality controlled through visual inspection of gray and white matter boundaries, and manually corrected where appropriate.

Diffusion-weighted MRI was corrected for eddy currents, movement artifacts, and magnetic field inhomogeneities using FSL top-up and eddy tools^35^. We applied generalized q-sampling imaging reconstruction in DSI Studio with a diffusion sampling length ratio of 1.25, followed by deterministic tractography^36^. Tractography generated approximately 2 million tracts per subject, with tracking parameters configured as follows: Runge-Kutta method with a step size of 1 mm, whole-brain seeding, initial propagation direction set to all fiber orientations, minimum tract length of 15 mm, maximum tract length of 300 mm, and topology-informed pruning applied with one iteration to remove false connections. Linear registration was applied to transform the tracts generated in the diffusion space to the preoperative T1-weighted MRI space.

### Segmentation of the Piriform

To segment the piriform we leveraged a recently published dataset of manually segmented left and right piriform cortices in 30 healthy controls^37^. Using the manually segmented T1w images from these 30 subjects, we trained an Automated Segmentation of Hippocampal Subfields (ASHS)^38^ model for piriform segmentation. An ASHS model consists of a combination of joint label fusion (JLF)^39^ and corrective learning (CL)^40^ for segmenting unseen data based on a manually labeled atlas set. All of this is performed using the open-source ASHS codebase (https://sites.google.com/view/ashs-dox). While originally developed for hippocampal subfield segmentation, this multi-atlas segmentation approach can be adapted to segment other structures. After training, the T1w images from the Penn, Emory, and HCP subjects were segmented using the pre-trained piriform atlas package. We will share our pre-trained ASHS package for piriform segmentation as part of the distributed segmentation service of ITK-SNAP (https://dss.itksnap.org/) as well as a downloadable package upon publication of the manuscript.

### Functional Gradient Analysis

To better understand the location of the piriform cortex in the functional manifold of adjacent structures, we performed a functional gradient analysis of the piriform. Functional gradients provide an overview of the similarity in functional connectivity across brain structures by performing a non-linear embedding of each voxel in a manifold where voxels with similar functional connectivity end up closer together. We performed our functional gradient analysis in the hippocampus, amygdala, thalamus, putamen, pallidum, caudate (from the Harvard-Oxford subcortical atlas) and piriform (averaged from all subjects in MNI space), using the connectivity of these structures to cortical gray matter as the input to a laplacian embedding algorithm^20^.

### Seed-to-voxel Functional Connectivity

To study the functional profile of the piriform cortex in controls and TLE, we calculated the seed-to-voxel connectivity profile for the left and right piriform following our previous approach^21^ (**Figure 1**). We used each subject’s individual piriform segmentation, registered to MNI space, as the piriform ROI. For each subject, and each ROI (left piriform and right piriform), we estimated the temporal average of the BOLD signal within the ROI, and computed the temporal Pearson correlation between the average ROI signal, and every other voxel in the brain. This resulted in volumetric seedmaps representing the functional connectivity between the piriform and every other voxel in the brain.

**Figure 1.**
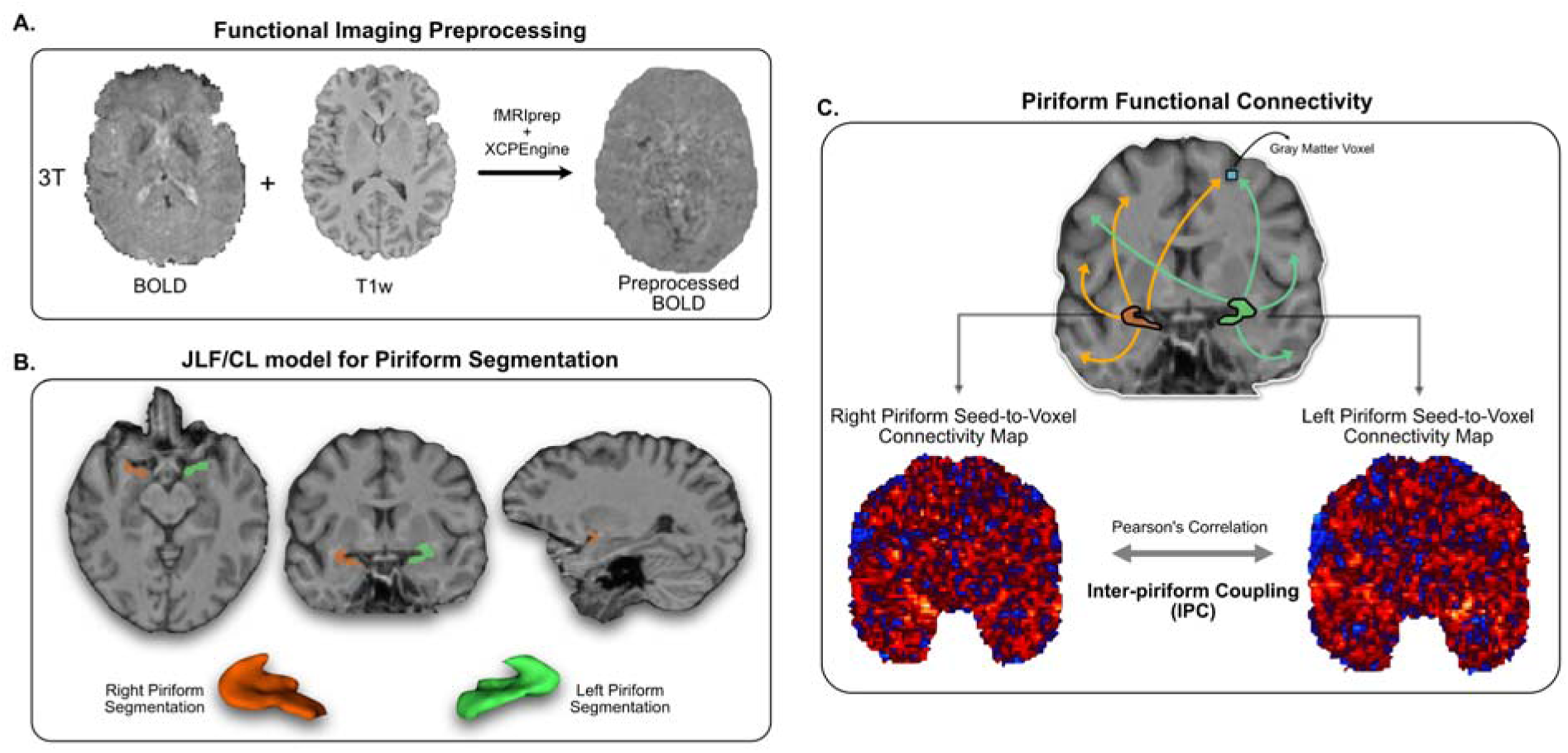
Methodological Overview: **Panel A.** We preprocessed resting-state fMRI data with fMRIprep and XCPEngine. **Panel B.** We trained a joint-label fusion/corrective learning based algorithm to automatically segment the piriform cortex from T1w weighted MRIs. We show an example segmentation generated by our trained algorithm, as well as a volumetric rendering, of the left and right piriform cortices from a control subject. **Panel C.** Using the preprocessed rs-fMRI data, and the piriform segmentations, we then compute seed-to-voxel connectivity maps, where the average temporal signal in the piriform segmentation is correlated with every voxel in the brain. We show two example seed-to-voxel maps, one for the left piriform and one for the right piriform. Correlation between these two maps, after smoothing and gray matter voxel thresholding, generates the inter-piriform coupling (IPC) metric we use throughout the manuscript.

We harmonized these seedmaps using NeuroCombat to ensure consistency across acquisition protocols^41^. Briefly, we vectorized each seedmap into a 1-dimensional vector for each subject. We then created a 2-dimensional dataset of [seedmaps X subjects] and applied NeuroCombat with a batch indicator variable corresponding to each acquisition site. We included an indicator variable for TLE and HCs as a covariate to preserve in the model, to ensure group differences were not disrupted by harmonization. After harmonization, we reshaped the seedmaps back to a 3-dimensional configuration. We zeroed negative connections to ensure that increases or decreases in connectivity could be interpreted as such, and smoothed the connectivity seedmaps with a Gaussian kernel of σ = 2.5.

### Inter-piriform Coupling

The piriform cortex is a bilateral structure, with structural connections and functional activity spanning both hemispheres^28–30^. Therefore, we hypothesized that differences in connectivity between piriform cortices might indicate functional reorganization in epilepsy. To quantify this reorganization, we measured the inter-piriform coupling (IPC) as the correlation of functional connectivity across the gray matter between left and right piriform seed maps. For each subject’s piriform seedmap in MNI space, we used a cortical and subcortical gray matter mask (TemplateFlow: tpl-MNI152NLin2009cAsym_res-02_label-GM_probseg.nii, resliced to seedmap space)^42^ to identify gray matter voxels, and calculated Spearman’s rank correlation between the values from the left and right piriform seed maps (**Figure 1C**).

### Whole-brain Integration and Segregation

We explored whether differences in IPC could be associated with global functional network properties of integration and segregation, as we have previously shown that these properties are associated with epilepsy subtype and surgical outcomes^31^. To measure differences in integration and segregation between groups, we computed the small-worldness of the whole brain network, as defined by Humphries and Gurney^43^. A more segregated network will have a lower small-worldness than a more integrated network. Briefly, we used the Harvard-Oxford cortical and subcortical atlas to define brain parcellations. We then estimated the absolute value of the pairwise Pearson correlation of the preprocessed BOLD signal between brain parcels to generate whole-brain functional connectivity matrices. We then applied density-based thresholding to generate binary networks at densities ranging from 0.1 to 0.5 in steps of 0.05^31^. At each density threshold, we estimated the small-worldness for each subject.

### Tractography of the Piriform

To estimate the structural integrity of the tracts emanating from the piriform across subgroups, we identified the streamlines connecting each piriform and every ROI in native space in the Desikan-Killiany-Tourvelle (DKT) atlas with an end-to-end tracking approach. We measured fractional anisotropy (FA) along each tract, and mean FA across all tracts connecting the two regions as a measure of structural connectivity. Because it is not always possible to identify tracts connecting two brain regions for any given subject, we pooled the average of the mean FA across all subjects of a given group within a brain region. Then for each region, we computed the difference between this metric for each group (e.g. controls vs. TLE). Positive values would indicate higher mean FA for the first group for that specific brain region, and vice versa.

### Shape Analysis of the Anterior Commissure

We identified the anterior commissure as the main bundle connecting both piriform cortices. We performed tractographic shape analysis of the anterior commissure using DSI-Studio AutoTrack^44,45^ as a way to measure differences in the structural basis of IPC. Details of this procedure are available in the supplementary methods.

### Statistical Analysis

We performed comparisons of individual metrics (e.g. piriform volumes, IPC differences, etc.) between TLE subgroups and HCs using two-sample, two-tailed t-tests, corrected for multiple comparisons using a Bonferroni procedure (*p_BON_*). We used Cohen’s *d* as an estimate of effect size considering effect sizes between *d*=0.5-0.8 as medium size, and *d*>0.8 as large effects^46^. We report the absolute value of Cohen’s *d*.

To compare functional seedmaps, we applied voxel-wise analyses using Threshold Free Cluster Enhancement (TFCE)^47^ with FSL *randomise*^48^, either in a one-sample (e.g. when comparing hippocampal and piriform seedmaps for the same subjects), or in a two-sample (e.g. when comparing seedmaps from TLE subgroups with controls) t-test. We used 5000 permutations and a threshold of *p*<0.05 to determine statistical significance. We limited our voxelwise analyses to cortical and subcortical gray matter voxels since fMRI preprocessing removed CSF and white matter signal. For two-sample voxel-wise comparisons between TLE subgroups, and between TLE subgroups and controls, we included age as an additional covariate in our design matrix.

## Results

### Automated segmentation of the piriform cortex

We used a multi-stage segmentation approach based on ASHS for segmenting the piriform cortex (**Figure 1A,B**). We found in a leave-one-out cross-validation (LOO-CV) no difference (left piriform: *p_BON_*=0.99, Cohen’s *d*=0.09; right piriform: *p_BON_*=0.66, Cohen’s *d*=0.23) between the manually segmented piriform volumes from the training set (left piriform: 460.75 mm^3^, 95% CI: [422.68, 498.83]; right piriform: 452.52 mm^3^, 95% CI: [456.96, 514.07]), and the automatically segmented piriform volumes (left piriform: 452.52 mm^3^, 95% CI: [426.92, 478.22]; right piriform: 467.12 mm^3^, 95% CI: [493.06, 495.18]) (**Supplementary Figure 1**). We also found good spatial overlap between the manual piriform segmentations in the training set and the LOO-CV automatic segmentations, with Dice coefficients of 0.67 (95% CI: [0.65, 0.70]) for the left piriform and 0.70 (95% CI: [0.68, 0.72]) for the right piriform. We note that the piriform is a small structure, therefore differences in segmentation of a few voxels can have a drastic impact on the Dice score^49^.

### Piriform volumetry does not lateralize the seizure-onset zone

We first investigated if the volume of the piriform cortices varied depending on the lateralization of the seizure onset zone. Consistent with previous studies^37,50^, we were unable to conclusively identify seizure-onset zone lateralizing findings based upon piriform volumetry (**Supplementary Figure 1**), with similar left and right piriform volumes in L-TLE (left piriform: 516.46 mm^3^, 95% CI: [491.87, 541.06]; right piriform: 509.64 mm^3^, 95% CI: [491.49, 527.80]; paired t-test, *p_BON_*=0.99, Cohen’s *d*=0.10) and R-TLE (left piriform: 514.74 mm^3^, 95% CI: [486.74, 542.74]; right piriform: 528.62 mm^3^, 95% CI: [492.44, 564.80]); paired t-test, *p_BON_*=0.99, Cohen’s *d*=0.18). We found similar values in healthy controls (left piriform: 505.96, 95% CI: [481.16, 530.77]); right piriform: 483.98 mm^3^, 95% CI: [455.00, 512.96]; paired t-test, *p_BON_*=0.79, Cohen’s *d*=0.31). This is in contrast to the lateralizing findings seen in the hippocampus (**Supplementary Figure 2**).

### The piriform of healthy controls is functionally connected to the ventral attention network

Using a voxel-wise procedure in control subjects, we measured the regions to which the piriform is highly connected to, relative to the hippocampus. We use the hippocampus as a reference as this is the most commonly studied epileptogenic structure, allowing us to better understand the unique contribution of the piriform to seizure networks. We found that the piriform is highly connected (TFCE, p<0.05) to the insula, putamen, and the opercular cortex (**Figure 2A**). A complete list of brain regions of connectivity is shown in **Supplementary Tables 3-10**. This spatial pattern of connectivity resembles that of the salience ventral attention network from Yeo and Krienen et. al.^51^ (**Figure 2B**), with a Dice overlap between significant voxels and the Yeo-Krienen ventral attention network of 0.25 for the left piriform network, and 0.15 for the right piriform network. This is in contrast to the areas of increased connectivity in the hippocampus, where the default mode network is the predominant canonical network, with Dice overlaps of 0.38 and 0.23 for the left and right hippocampus respectively. This shows that despite their close proximity, the hippocampus and the piriform, both regions known to be involved in the epileptogenicity of TLE, have different functional connectivity profiles. These findings were reproduced in a set of subjects from the Human Connectome Project (**Supplementary Figure 3**)

**Figure 2.**
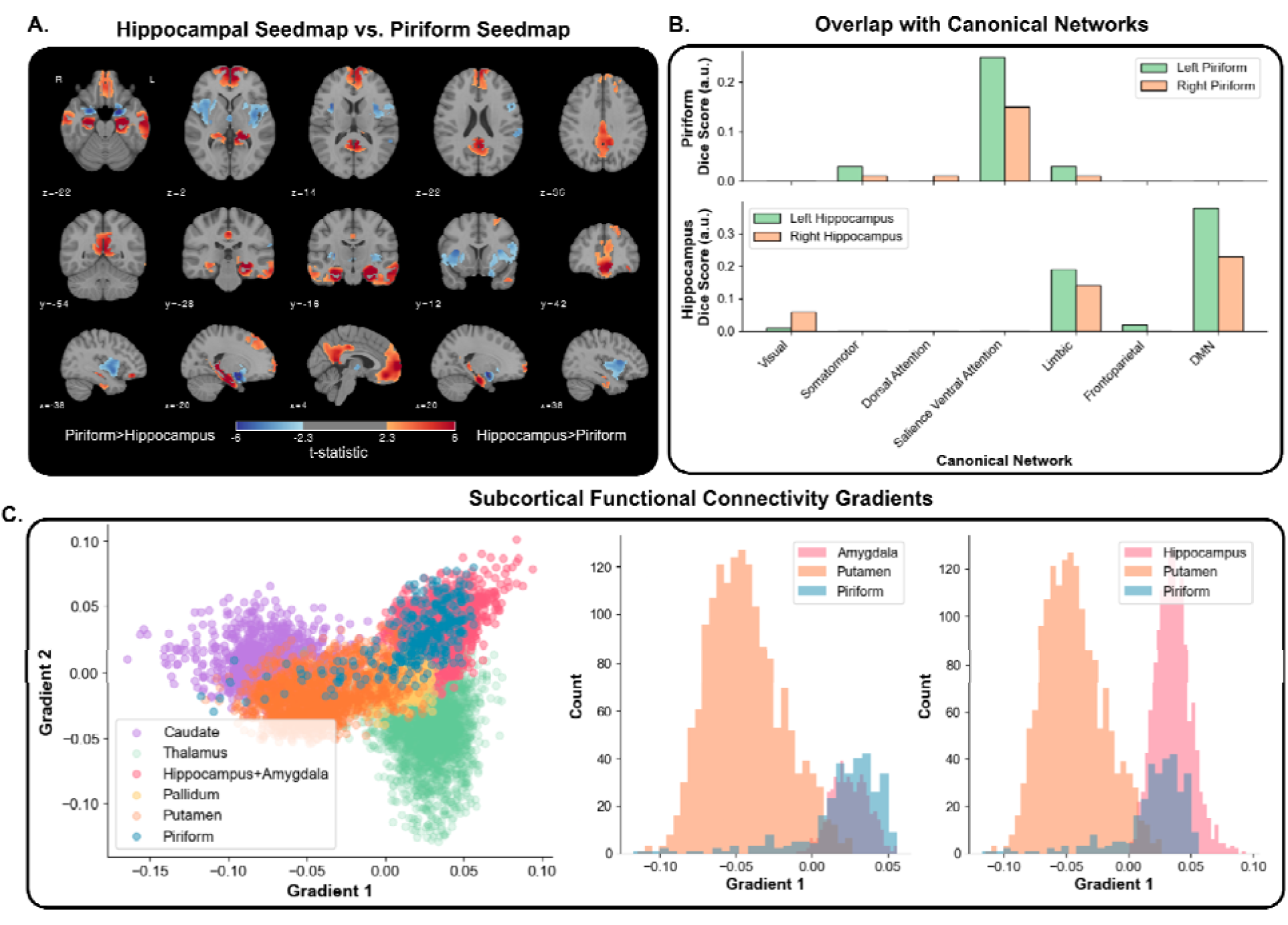
Normative piriform connectivity: **Panel A.** demonstrates the voxelwise comparison between the piriform and the hippocampal seedmaps in healthy control subjects. Shown voxels survived threshold-free cluster enhancement significance testing (p<0.05), and the colormap represents the corresponding t-statistic of each voxel. We show the results for the left piriform. **Panel B.** shows the Dice overlap score between the normative piriform mask (top) and the normative hippocampal mask (bottom) across the Yeo-Krienen canonical networks. **Panel C.** shows the mean functional connectivity gradients of subcortical structures and the piriform across healthy control subjects. In the scatterplot (left) each point represents a voxel. The histograms (middle and right) show the projection of the scatterplot along the Gradient 1 axis (x-axis).

Functional gradients provide a global overview of the connectivity pattern across brain structures. Placing the piriform in the principal subcortical gradient provides a deeper understanding of the connectivity of this structure to the rest of the brain, relative to the amygdala, hippocampus and other subcortical structures. While overlapping with the hippocampus and amygdala (**Figure 2C**), the piriform (mean principal gradient: 0.020, 95% CI:[0.016,0.023]) still has a significant offset in the principal gradient relative to the amygdala (mean principal gradient: 0.025, 95% CI:[0.024,0.026]; *p_BON_*<0.001, Cohen’s *d*=0.24) and hippocampus (mean principal gradient: 0.036, 95% CI:[0.035,0.036]; *p_BON_*<0.001, Cohen’s *d*=0.81), driven by a longer tail that overlaps with the pallidum and putamen. This places the piriform, functionally, as a transitional structure from the basal ganglia into the amygdala and hippocampus.

### Inter-piriform coupling identifies subgroups of TLE patients and their outcomes

The piriform cortex is a bilateral structure, with structural connections and functional activity spanning both hemispheres^28–30^. Therefore, we hypothesized that differences in connectivity between piriform cortices might indicate functional reorganization in epilepsy. To quantify this reorganization, we measured the inter-piriform coupling (IPC) as the correlation of functional connectivity across the gray matter between left and right piriform seedmaps. We found that the IPC was significantly lower for TLE participants (0.40, 95% CI: [0.32, 0.48]) than it was for healthy control participants (0.54, 95% CI: [0.45, 0.64], Cohen’s *d =* 0.53, *p_BON_* = 0.030; **Figure 3A**). We did not find this difference to be associated with the laterality of epilepsy (**Supplementary Table 1; Supplementary Figure 4**). We did find, however, that this difference was driven by a subset of TLE participants that had a much lower IPC than the rest. A disproportionate number of TLE participants (19/55) had an IPC below 0.20 compared to healthy controls (2/26) (Chi-Square = 6.62, p=0.010). This value of 0.20 is 1.5 standard deviations away from the mean IPC of controls. We defined this subgroup of TLE patients as Low-IPC TLE participants, to distinguish them from the High-IPC TLE participants. We repeated this analysis with inter-hippocampal coupling (**Supplementary Figure 5**), and found it also to be significantly lower in TLE (0.43, 95% CI: [0.35, 0.52]) than in healthy controls (0.68, 95% CI: [0.62, 0.74]; Cohen’s *d* = 0.90, *p*<0.001), but unlike the IPC, the hippocampal coupling had a normal distribution in the TLE subgroup, and both low and high-IPC subgroups had reductions in inter-hippocampal coupling (**Supplementary Figure 5**).

**Figure 3.**
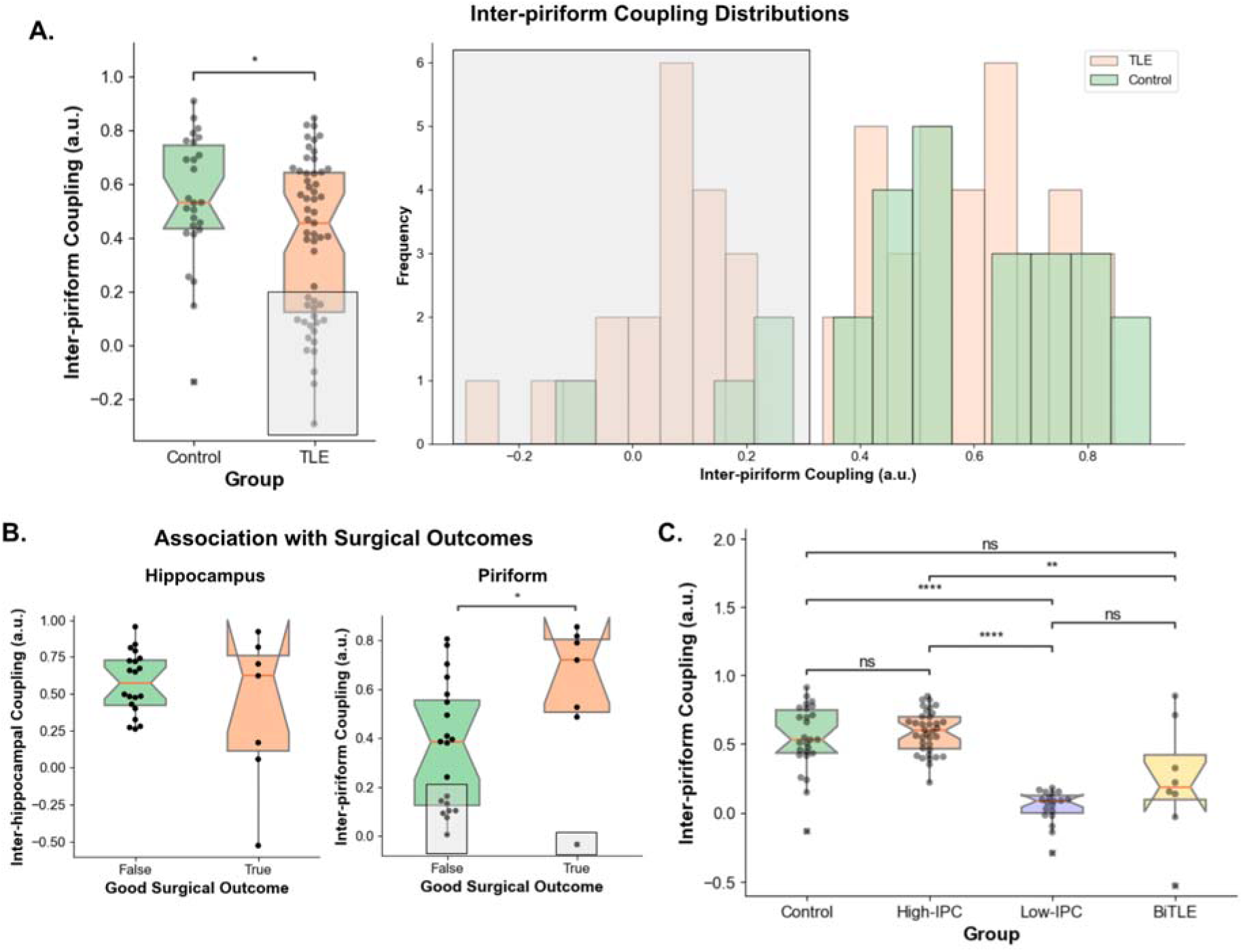
Inter-piriform Coupling: **Panel A.** shows the inter-piriform coupling (IPC) values between the healthy control subjects and the TLE subjects. The gray box represents the subjects assigned to the Low-IPC group based on the value of their IPC. **Panel B.** shows the value of the inter-hippocampal coupling (left) and the IPC (right) between TLE subjects with good (Engel IA) and poor (Engel IB-IV) surgical outcomes. **Panel C.** shows the IPC across control subjects, High-IPC, Low-IPC and bilateral TLE (BiTLE) subjects. n.s. - not significant. *p<0.05, **p<0.01, ***p<0.001, ****p<0.0001. *p-* values are Bonferroni corrected where appropriate.

Measuring the association between IPC and surgical seizure outcomes at 24 months (**Figure 3B**), we found that participants that were seizure free (Engel IA) had a higher IPC (0.60, 95% CI: [0.32, 0.88]), than participants that were not seizure free (Engel IB-IV) (0.36, 95% CI: [0.23, 0.48]; Cohen’s *d* = 0.94, p = 0.05). These findings demonstrate that low IPC is a sensitive, but not specific, measure of poor surgical outcome at 24 months post-surgery (sensitivity: 85.71%, 95% CI: [19.12%, 99.64%]; specificity: 40.00%, 95% CI: [19.12%, 63.95%]). A smaller effect in the same direction was also present at 12 months post-surgery (Cohen’s *d*=0.48, p=0.24; **Supplementary Figure 4**). Performing the same coupling measurement, but between hippocampal seedmaps, we did not find a significant effect between seizure free and non-seizure free groups (**Figure 3B**).

Comparing demographic characteristics between the Low-IPC and High-IPC groups, we did not find a significant difference in the distribution of gender, acquisition protocols, or left and right TLE participants (**Supplementary Table 1**) between the two subgroups (Chi-squared test p>0.05). We did find that the Low-IPC group had slightly older participants (43.17 years, 95% CI: [38.08, 48.25]) than the High-IPC groups (38.52 years, 95% CI: [34.65, 42.40]), but this difference was not significant (Cohen’s *d* = 0.42, *p_BON_* = 0.277). However, both low (Cohen’s *d* = 1.25, *p_BON_*< 0.001) and high (Cohen’s *d* = 0.75, *p_BON_*= 0.008) IPC subgroups had participants older than the control participants (30.38 years, 95% CI: [26.20, 34.55]). We also found a weak negative correlation between IPC and age (Spearman’s *r* = −0.28, *p*=0.011). To test the effect of age on IPC differences, we regressed age from our IPC measurement, but differences in IPC between subgroups remained significant after regression (**Supplementary Figure 6**). We also tested whether piriform segmentation and MNI registration differences or BOLD signal factors were contributing to the differences in IPC between groups. We found no difference in piriform segmentation volumetry, piriform centroid coordinates after MNI registration, or BOLD signal temporal signal-to-noise ratio (tSNR) between Low-IPC, High-IPC and control groups (**Supplementary Figure 7**). We additionally tested whether the direct ROI-to-ROI connectivity between left and right piriform, measured directly in native space, was lower for the Low-IPC group than the High-IPC and control groups, and found this to be the case (**Supplementary Figure 7**), with this measurement and IPC being highly correlated (Spearman’s *r* = 0.67, p<0.001). We were able to replicate the presence of a Low-IPC subgroup in an external dataset of TLE patients (**Supplementary Figure 8**), although the separation between low and high IPC in this dataset appeared at IPC=0.30.

### The piriform has altered connectivity to its normative network in TLE

To better understand the connections driving the differences between low and high IPC groups, we performed a voxel-wise comparison between Low-IPC, High-IPC and control subjects for the left and right piriform seed maps. To ensure that the differences found were not driven by age differences between the groups, we included age as a covariate in the design matrix of our analysis. We found that relative to controls, the left piriform in the Low-IPC group had right-sided (contralateral) disconnectivity (TFCE, p<0.05) to the posterior insula, the right piriform, and the planum temporale and parietal opercular cortex junction (**Figure 4A**). A similar disconnectivity is seen for the Low-IPC group relative to the High-IPC group, although the pattern is more widespread, and also involves some left-sided (ipsilateral) disconnectivity from the insula (**Figure 4B**). No differences in connectivity were seen in the right piriform seedmaps for the Low-IPC group. For the High-IPC group relative to controls, we see the opposite effect. We found that both the left (**Figure 4C**) and right (**Figure 4D**) piriform had increased connectivity (TFCE, p<0.05) to the left anterior insula, a key node in the salience attention network.

**Figure 4.**
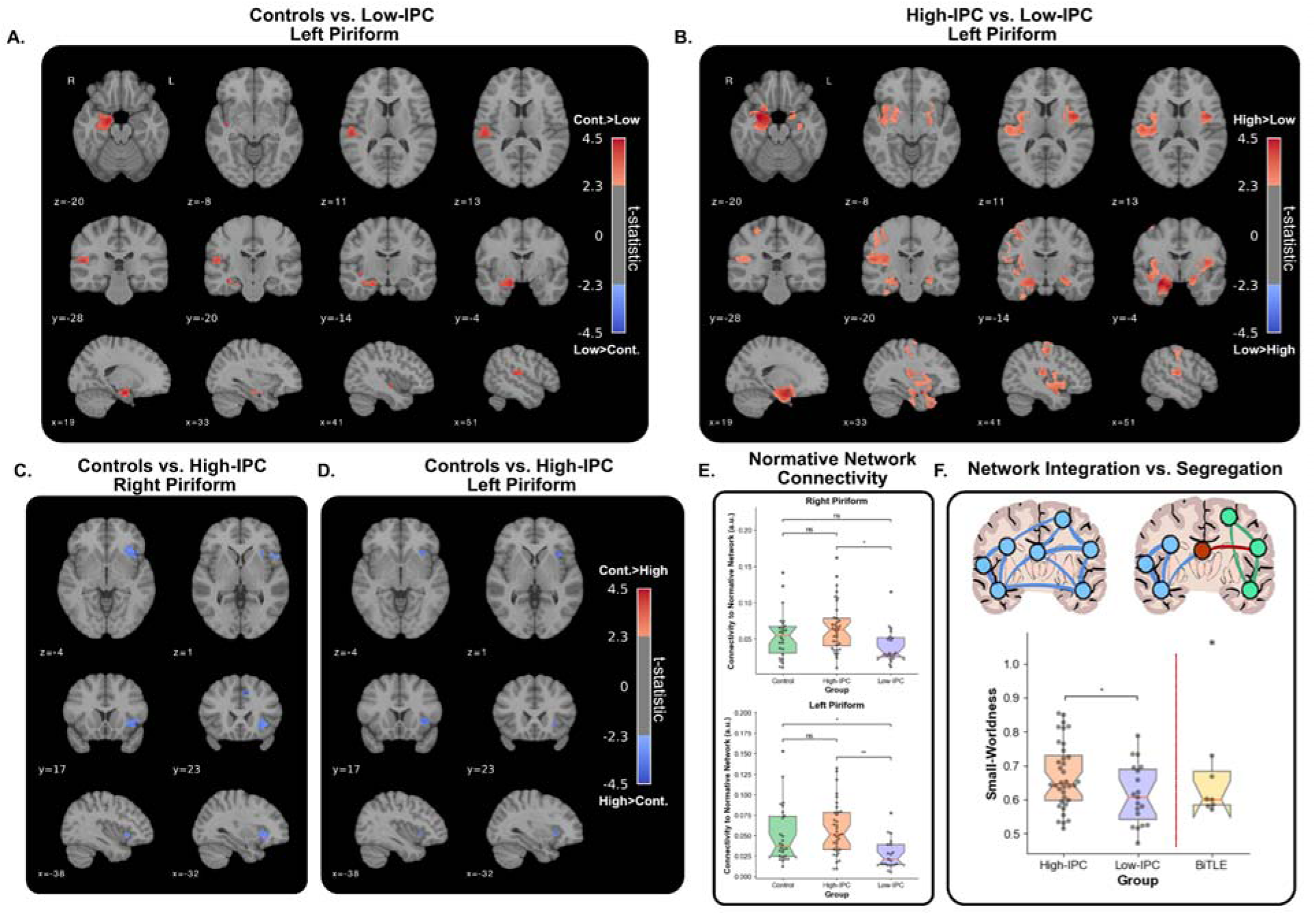
Functional connectivity differences based on inter-piriform coupling: **Panel A.** shows the resulting voxelwise comparison between the left piriform seedmaps of healthy control subjects and Low-IPC TLE subjects. **Panel B.** the voxelwise comparison between the left piriform seedmap in High-IPC and Low-IPC TLE subjects. **Panels C.** and **D.** show the voxelwise comparison between the right (**C.**) and left (**D.**) piriform seedmaps between control subjects and the High-IPC group. For panels **A-D.**, shown voxels survived threshold-free cluster enhancement significance testing (p<0.05), and the colormap represents the corresponding t-statistic of each voxel. **Panel E.** shows the mean value of each seedmap inside the normative mask generated in Figure 2, representing the connectivity of each piriform (top - left piriform; bottom - right piriform) in each subgroup to their normative network. **Panel F.** shows the small-worldness, as a measure of network integration, between the high and low IPC TLE subjects. The bilateral TLE (BiTLE) cohort is shown as a reference to demonstrate its similarity with the Low-IPC group. n.s. - not significant. *p<0.05, **p<0.01, ***p<0.001, ****p<0.0001. *p-*values are Bonferroni corrected where appropriate.

The spatial pattern of hyperconnectivity in High-IPC and disconnectivity in Low-IPC follows the normative piriform network we uncovered in healthy controls. To test whether the spatial pattern of connectivity and disconnectivity is within the normative piriform network, we generated a normative network mask (**Figure 2A**) by selecting voxels with significantly higher (TFCE, p<0.05) connectivity to the piriform than the hippocampus. We then measured the mean value of the left and right piriform seedmaps within this mask, for each subject. We indeed found that the Low-IPC group had significantly lower connectivity to the normative piriform network for both left (0.0278, 95% CI: [0.0146, 0.0288]) and right (0.0406, 95% CI: [0.0294, 0.0517]) piriform, compared to the High-IPC group (left piriform: 0.0577, 95% CI: [0.0473, 0.0681], Cohen’s *d* = 1.08, *p_BON_* = 0.001; right piriform: 0.0660, 95% CI: [0.0549, 0.0772], Cohen’s *d* = 0.83, *p_BON_* = 0.017). Control subjects had normative piriform connectivities between the values of the Low and High-IPC groups (left piriform: 0.0506, 95% CI: [0.0371, 0.0640]; right piriform: 0.0547, 95% CI: [0.0423, 0.0670]), statistically higher from those of Low-IPC group in the left piriform (Cohen’s *d* = 0.79, *p_BON_* = 0.040). We were able to replicate these findings in our external validation cohort with the mask derived from our primary cohort (**Supplementary Figure 8**), as well as in our primary cohort with a mask derived from HCP subjects (**Supplementary Figure 3**).

These findings demonstrate that while the Low-IPC has disconnectivity from the normative piriform network, the High-IPC group has hyperconnectivity to the normative piriform network. This finding is relevant for the High-IPC group, because even though we observe that the IPC values of the High-IPC group are in the same range as those from controls (**Figure 3C**), we still observe increased connectivity relative to controls in this subgroup, suggesting that both low and high-IPC TLE participants have altered connectivity relative to healthy controls.

### Low inter-piriform coupling suggests increased network segregation

The pattern of disconnectivity in the Low-IPC group and hyperconnectivity in the High-IPC group could be associated with high and low whole-brain segregation. In the context of brain networks, a highly integrated network requires a small number of connections for a signal to travel between any two nodes in the network. The opposite, a highly segregated network, requires many connections. We previously demonstrated that low segregation was associated with poor outcome and bilateral disease in TLE^31^. Given our prior work, and our current findings, we hypothesized that the Low-IPC group would have decreased whole-brain network segregation, relative to the High-IPC group. We measured network segregation using small-worldness^43,52^, with lower small-worldness representing increased segregation.

We found that at network densities around 0.30, the whole-brain small-worldness between low and high-IPC groups began to diverge, with the Low-IPC group having lower small-worldness (**Supplementary Figure 9**). Since a small-world network by definition would have a small-worldness > 1, and brains are small-world networks^53^, we compared the small-worldness between low and high IPC groups at a density threshold of 0.45, which would ensure all subjects would have a small-worldness above 1. At this density threshold (**Figure 4F**), we found that the Low-IPC group had lower small-worldness (1.07, 95% CI: [1.04, 1.09]) than the High-IPC group (1.11, 95% CI: [1.08, 1.13]; Cohen’s *d* = 0.60, *p*=0.043 (uncorrected)). Consistent with our prior work, we also found that bilateral TLE (BiTLE; demographics in **Supplementary Table 1**) subjects had a similar small-worldness to the Low-IPC group (1.08, 95% CI: [1.01, 1.15]; Cohen’s *d* = 0.20, *p*=0.66 (uncorrected)). As with our prior work, we also found that control subjects have segregation comparable to the BiTLE group (1.07, 95% CI: [1.06, 1.09]; not plotted). Finally, to relate the small-worldness seen in BiTLE to the IPC, we computed the IPC in the BiTLE subgroup (0.23, 95% CI: [-0.07, 0.52]; **Figure 3C**), and found that this value was slightly higher but statistically equivalent to that of the Low-IPC group (0.04, 95% CI: [-0.01, 0.10]; Cohen’s *d* = 0.79, *p_BON_* = 0.54), but significantly lower than for the High-IPC group (0.58, 95% CI: [0.53, 0.63]; Cohen’s *d* = 1.67, *p_BON_* = 0.001).

Together, these findings support the idea of Low-IPC corresponding with low integration, and consequently high segregation, as demonstrated by the lower small-worldness and this group’s similarity to BiTLE participants. This suggests that while the original low and high IPC subgroups were defined based on the values measured through piriform coupling, these subgroups have whole brain network differences that are independent of values measured directly in the piriform, hinting at a larger network involvement.

### Low inter-piriform coupling is associated with tractographic changes

To uncover whether structural connectivity can elucidate a mechanism for low and high IPC, we quantified the fractional anisotropy for white matter tracts originating from the piriform and terminating in other cortical and subcortical regions (**Figure 5A**). We found that the mean FA for ipsilateral piriform connections (**Figure 5C**) was lower in the Low-IPC (mean difference: 0.035, 95% CI: [0.0246, 0.0453], one-sample t-test *p_BON_*<0.001), and High-IPC (mean difference: 0.027, 95% CI: [0.0163, 0.0378], one-sample t-test *p_BON_*<0.001) groups than in healthy controls. For contralateral piriform connections (**Figure 5C**), the mean FA was also lower for the Low-IPC group (mean difference: 0.0267, 95% CI: [0.0189, 0.0346], one-sample t-test *p_BON_*<0.001) and the High-IPC group (mean difference: 0.0083, 95% CI: [0.0021, 0.0145], one-sample t-test *p_BON_*=0.043), but the difference was higher for the Low-IPC group (Cohen’s *d* = 0.58, *p_BON_*=0.001). These results suggest that both the low and high IPC groups have a similar degree of impaired ipsilateral piriform structural connectivity, but the Low-IPC group has higher impairment in contralateral piriform structural connectivity. This contralateral effect in the Low-IPC group is consistent with the contralateral findings we observed in the voxelwise functional connectivity analysis, suggesting a possible structural basis for our functional findings.

**Figure 5.**
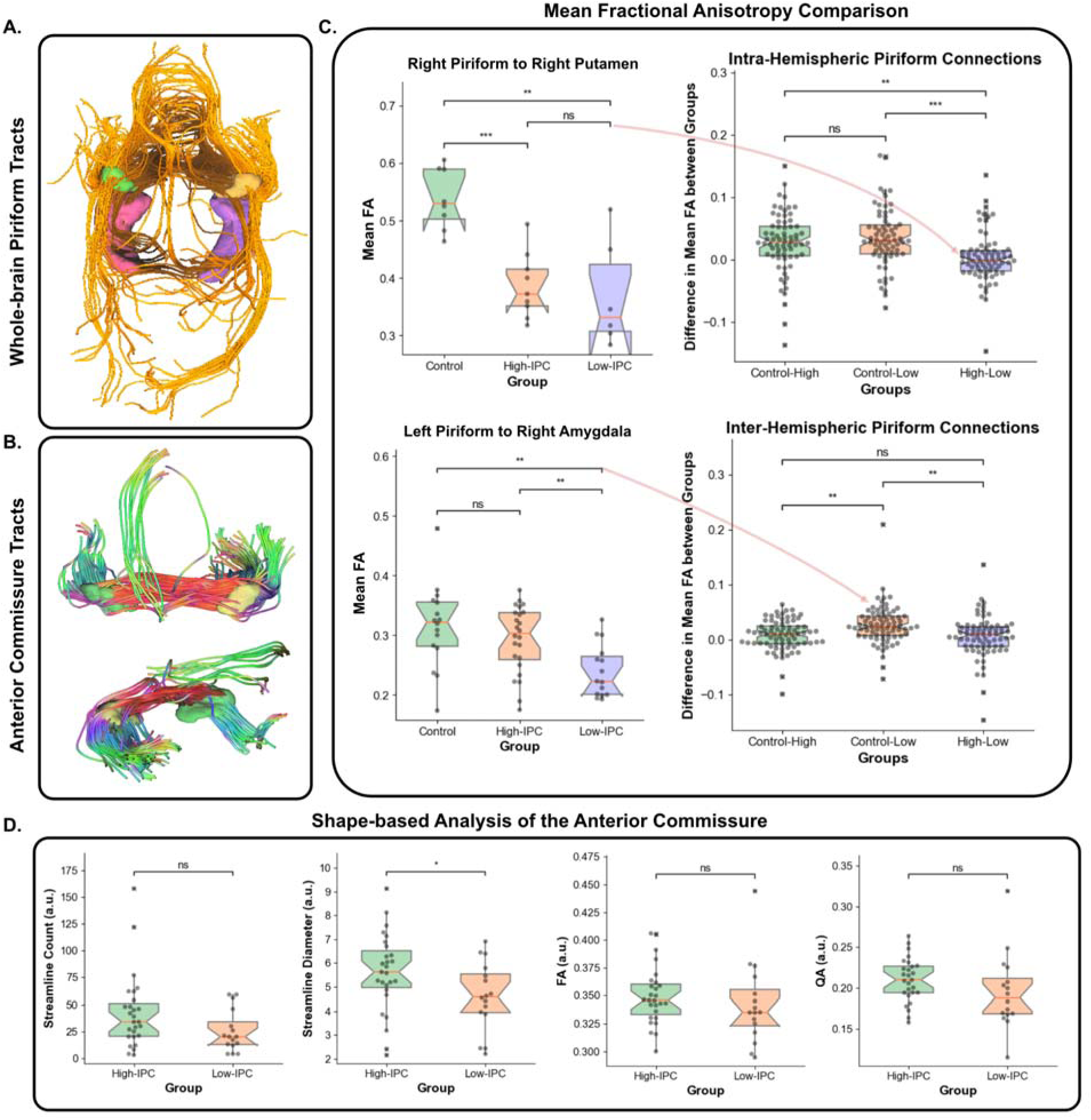
Structural connectivity differences based on inter-piriform coupling: **Panel A.** shows an example of the tracts generated by DSI Studio autotrack after defining the left piriform (shown in yellow) as a seed and the right piriform (shown in green) as pass-through ROI. Both hippocampi are shown in purple (left hippocampus) and pink (right hippocampus). **Panel B.** shows an example of the tracts identified as belonging to the anterior commissure for a representative subject. The left and right piriform segmentations are overlaid on top of the anterior commissure tracts for visualization purposes. **Panel C.** shows the comparison of the mean fractional anisotropy between the piriform and other brain regions in the DKT atlas. Each point on the boxplots to the right represents the difference in mean FA between two groups for a brain region. Examples of these brain regions are shown on the left of the boxplot, with the red arrows showing the corresponding point belonging to that region. The top row shows intra-hemispheric piriform connections (i.e. left to left, right to right) and the bottom row shows inter-hemispheric piriform connections (i.e. left to right, right to left). **Panel D.** shows the comparison of tract-derived shape-based metrics for the anterior commissure between the high and low-IPC groups. n.s. - not significant. *p<0.05, **p<0.01, ***p<0.001, ****p<0.0001. *p-*values are Bonferroni corrected where appropriate.

As expected based on prior neuroanatomical studies^54^, the main tract connecting the two piriform regions was the anterior commissure (**Figure 5B**), therefore we hypothesized that differences in this tract would be associated with differences in IPC. We identified the anterior commissure in each subject, and quantified the streamline count, diameter, mean FA and quantitative anisotropy (QA) for this structure (**Figure 5D**). We found no differences in FA (Low-IPC: 0.34, (95% CI: [0.32, 0.36]; High-IPC: 0.35, 95% CI: [0.34, 0.36]; Cohen’s *d* = 0.25, *p*=0.45) or QA (Low-IPC: 0.20, 95% CI: [0.17, 0.22]; High-IPC: 0.21, 95% CI: [0.20, 0.22]; Cohen’s *d* = 0.42, *p*=0.20) between the Low-IPC and High-IPC groups. We did find, however, a significantly smaller streamline diameter in the Low-IPC (4.60, 95% CI: [3.90, 5.31]) than the High-IPC (5.61, 95% CI: [5.00, 6.23], Cohen’s *d* = 0.66, *p*=0.047) group, as well as a smaller streamline count for the Low-IPC (25.31, 95% CI: [15.82, 34.80]) than the High-IPC (42.44, 95% CI: [29.42, 55.47], Cohen’s *d* = 0.59, *p_BON_*=0.077) group. These findings suggest smaller and more sparse streamlines connect the two piriform through the anterior commissure in the Low-IPC group, than in the High-IPC group. This could be another potential structural basis for the Low-IPC in this subset of TLE patients. Furthermore, because the identification and characterization of the anterior commissure is independent of the piriform segmentation, this is another finding that reproduces group differences between the high and low IPC groups, while being independent of the IPC measurement itself.

## Discussion

In this work, we shed light on the structural and functional abnormalities that feature the piriform cortex as a key node within the seizure-onset zone network of TLE. Our findings show that the functional network of the piriform cortex aligns with the spatial distribution of the salience ventral attention network, contrasting sharply with the hyperconnectivity observed between the hippocampus and the default mode network. We also identified distinct subgroups among TLE patients based on inter-piriform coupling, revealing that lower IPC values are associated with poor surgical outcomes, and we further showed that differences in IPC were driven by disconnectivity and hyperconnectivity to the normative piriform network. These connectivity abnormalities are in turn related to whole brain integration and segregation properties, hinting at a larger network involvement driving these changes. Finally, tractography provided evidence of a structural basis for these functional changes, showing decreased connectivity from the piriform to the rest of the brain in the presence of low IPC.

### Piriform as part of the epileptogenic network

We found that the piriform has high connectivity to the salience ventral attention network (SAN). The SAN is known to be heavily involved in visceroautonomic integration, likely due to its association with the anterior insula^55,56^. The relationship between the piriform cortex and the anterior insula underscores the integration of smell and taste processing. This is particularly relevant in the context of TLE, where both olfactory auras and visceral epigastric symptoms are common seizure manifestations^57^. The piriform cortex’s involvement in seizures may contribute to these autonomic symptoms. Furthermore, the piriform’s proximity to the amygdala, a region involved in processing emotions and autonomic functions, make this also a likely explanation.

Our results also shed some light as to what could be driving the decreased IPC in this cohort of TLE patients. We did not find differences in piriform volumetry between the low-IPC and high-IPC groups, therefore atrophy of the piriform is likely not a driver (as it can be with hippocampal atrophy). These differences could be due to neuroplastic changes, reorganizing the piriform’s connectivity to adapt to epileptogenic conditions. Such neuroplastic alterations could limit the piriform’s efferent connections to other brain regions, contributing to the observed differences in IPC and structural connectivity, and impacting the overall network dynamics in TLE patients. Finally, it is also possible that the decreased IPC is an epiphenomenon of the underlying brain injury or genetic abnormality associated with epilepsy.

### TLE phenotypes and implications for surgery

Our results suggest two different phenotypes driven by low and high inter-piriform coupling. The piriform is an inherently bilateral structure, therefore asymmetries in connectivity detected by decreased IPC are likely indicative of pathology. For the Low-IPC group, we found increased whole brain network segregation, as well as decreased piriform connectivity to its normative network contralaterally. From our prior work, we have seen increased network segregation being associated with worse outcomes and bilateral disease. We also found similar associations with decreased IPC. Overall, decreased IPC could be indicative of a segregated piriform network that is involved in epileptogenicity, but that might be functionally segregated from the main epileptogenic zone. Given the similarity of the Low-IPC group to BiTLE, it might also be indicative of bilateral involvement of the piriform, with the two piriform cortices acting as independent nodes in the SOZ. For this reason, even if the main SOZ is removed in individuals with low-IPC, the outcomes are poor if the piriform is not resected. Furthermore, if one piriform is resected in this group, because of the segregation of the two piriform cortices, it might not be sufficient to achieve a therapeutic effect, and both piriform cortices must be intervened upon to achieve a successful outcome. Unfortunately, few of the cases included in this study had their piriform removed after surgery (data not shown), making it impossible to directly assess these hypotheses with the available data.

The increased segregation of the piriform in the low-IPC group is also supported by decreases in structural connectivity from the piriform to the rest of the brain, which we found in the high-IPC group to an extent, but was more pronounced in contralateral connections for the low-IPC group. This is further supported by decreases in fiber diameter in the anterior commissure in the low-IPC group, suggesting decreased inter-piriform structural connectivity, and hence increased piriform segregation.

Our findings could have direct implications in the surgical management of TLE. Previous studies have demonstrated that incomplete resection of the piriform cortex leads to worse outcomes in epilepsy. Our findings suggest that while this might be true for a subset of TLE patients (low-IPC group), other TLE patients might still do well without removal of the piriform. As the high-IPC cohort has a more integrated brain network, removal of the main SOZ will directly influence piriform connectivity. It is also possible that for patients with high-IPC, neurostimulation of the piriform might be sufficient to control seizures more generally (due to higher integration), whereas in the low-IPC group, neurostimulation of a single piriform might not lead to seizure reduction due to the relative segregation of the network, potentially requiring bilateral neurostimulation to achieve a therapeutic target.

### Challenges and future directions

Our study has some limitations, notably in its primary focus on structural and functional connectivity without integrating metabolic or neurotransmitter-specific imaging data, which could unveil the neurobiological mechanisms underpinning the observed connectivity patterns. By incorporating positron emission tomography (PET) or magnetic resonance spectroscopy (MRS), we could enhance our understanding of the complex interplay between structural, functional, and biochemical changes in the piriform cortex. Future work could also leverage newer techniques such as glutamate-weighted chemical exchange saturation transfer (GluCEST) of the piriform, which at 7-Tesla field strengths has unveiled glutamatergic changes in the hippocampus of TLE patients^22^. Additionally, our analysis, rooted in cross-sectional data, limits our capacity to deduce causality or monitor the progression of connectivity alterations over time. Implementing longitudinal study designs with multiple follow-up assessments would allow us to observe changes in connectivity and their relationship with clinical outcomes over time. Consequently, we advocate for longitudinal research aimed at unraveling the temporal dynamics of piriform cortex connectivity alterations in epilepsy, focusing on how these changes evolve with disease progression or treatment interventions and their prognostic value.

## Conclusion

Our study of the piriform cortex’s role in temporal lobe epilepsy (TLE) presents novel insights into the functional and structural neural network alterations associated with this structure. We show that the piriform cortex exhibits differential connectivity patterns to the salience ventral attention network, with potential implications for understanding the neurobiological underpinnings of TLE, particularly in relation to seizure manifestations and surgical outcomes. The distinction between low and high IPC groups reveals variations in network integration and segregation, which are closely tied to the effectiveness of surgical interventions and potentially to the overall management of epilepsy. Our findings underscore the importance of considering the piriform cortex’s connectivity during surgical management, and future work is required to better understand how the piriform’s connectivity can be used to aid in therapy selection and management of drug resistant TLE.

## Data Availability Statement

Upon publication of our manuscript, we will provide the automated piriform segmentation algorithm as a tool for the research community through the free distributed segmentation service (DSS) of ITK-SNAP (https://dss.itksnap.org/). We will additionally provide a version of the Harvard-Oxford cortical and subcortical atlas that includes the piriform cortex. The code used to generate all analyses, figures, and statistical tests will be provided in a GitHub repository upon publication of the manuscript.

## Acknowledgments

AL and KAD received support from NINDS (R01NS116504). NS received support from American Epilepsy Society (953257) and NINDS (R01NS116504). EG was supported by awards UL1TR002378 and KL2TR002381 by the National Center for Advancing Translational Sciences (NCATS) of the National Institutes of Health (NIH). Emory data was part of the Connectome Abnormalities to Predict Epilepsy Surgery, obtained through NINDS R01NS110347.

## Author Contributions

**Alfredo Lucas:** Conceptualization, Methodology, Software, Validation, Formal analysis, Writing - Original draft preparation. **Marc Jaskir:** Writing - Review & Editing, Methodology. **Nishant Sinha:** Writing - Review & Editing, Methodology, Data Curation. **Akash Pattnaik:** Writing - Review & Editing, Methodology, Data Curation. **Sofia Mouchtaris:** Writing - Review & Editing, Methodology. **Mariam Josyula:** Data Curation, Writing - Review & Editing. **Nina Petillo:** Data Curation, Writing - Review & Editing. **Gulce N Dikecligil:** Writing - Review & Editing, Conceptualization. **Leonardo Bonilha:** Data Curation, Writing - Review & Editing. **Jay Gottfried:** Writing - Review & Editing, Conceptualization. **Ezequiel Gleichgerrcht:** Data Curation, Writing - Review & Editing. **Sandhitsu Das:** Data Curation, Conceptualization, Writing - Review & Editing. **Joel M. Stein:** Data Curation, Conceptualization, Methodology, Writing - Review & Editing. **James J. Gugger:** Conceptualization, Data Curation, Methodology, Writing - Review & Editing. **Kathryn A. Davis:** Supervision, Project administration, Funding acquisition, Data Curation, Conceptualization, Writing - Review & Editing.

