## Supplementary for "Connectivity of the Piriform Cortex and its Implications in Temporal Lobe Epilepsy"

**Supplementary Methods**

**Neuroimaging Processing**

Preprocessing was performed using fMRIPrep 20.2.3[1], which is based on Nipype 1.6.1[2].

*Anatomical data preprocessing*

For each subject, T1w images were corrected for intensity non-uniformity (INU) with N4BiasFieldCorrection[3], distributed with ANTs 2.3.3[4]. The T1w-reference was then skull-stripped with a Nipype implementation of the antsBrainExtraction.sh workflow (from ANTs), using OASIS30ANTs as target template. Brain tissue segmentation of cerebrospinal fluid (CSF), white-matter (WM) and gray-matter (GM) was performed on the brain-extracted T1w using FAST (FSL 5.0.9)[5]. A T1w-reference map was computed after registration of 2 T1w images (after INU-correction) using mri_robust_template (FreeSurfer 6.0.1)[6]. Brain surfaces were reconstructed using FreeSurfer's recon-all, and the brain mask estimated previously was refined with a custom variation of the method to reconcile ANTs-derived and FreeSurfer-derived segmentations of the cortical gray-matter of Mindboggle[7]. Volume-based spatial normalization to one standard space (MNI152NLin2009cAsym) was performed through nonlinear registration with antsRegistration (ANTs 2.3.3), using brain-extracted versions of both T1w reference and the T1w template. The following template was selected for spatial normalization: ICBM 152 Nonlinear Asymmetrical template version 2009c[8].

*Functional data preprocessing*

For the single BOLD run of each subject, the following preprocessing was performed. First, a reference volume and its skull-stripped version were generated using a custom methodology of fMRIPrep. A B0-nonuniformity map (or fieldmap) was estimated based on a phase-difference map calculated with a dual-echo GRE (gradient-recall echo) sequence, processed with a custom workflow of SDCFlows inspired by the epidewarp.fsl script and further improvements in HCP Pipelines[9]. The fieldmap was then co-registered to the target EPI (echo-planar imaging) reference run and converted to a displacements field map (amenable to registration tools such as ANTs) with FSL's fugue and other SDCflows tools. Based on the estimated susceptibility distortion, a corrected EPI (echo-planar imaging) reference was calculated for a more accurate co-registration with the anatomical reference. The BOLD reference was then co-registered to the T1w reference using bbregister (FreeSurfer) which implements boundary-based registration[10]. Co-registration was configured with six degrees of freedom. Head-motion parameters with respect to the BOLD reference (transformation matrices, and six corresponding rotation and translation parameters) are estimated before any spatiotemporal filtering using mcflirt (FSL 5.0.9)[11]. BOLD runs were slice-time corrected using 3dTshift from AFNI 20160207[12]. The BOLD time-series (including slice-timing correction when applied) were resampled onto their original, native space by applying a single, composite transform to correct for head-motion and susceptibility distortions. These resampled BOLD time-series will be referred to as preprocessed BOLD in original space, or just preprocessed BOLD. The

BOLD time-series were resampled into standard space, generating a preprocessed BOLD run in MNI152NLin2009cAsym space. First, a reference volume and its skull-stripped version were generated using a custom methodology of fMRIPrep. Several confounding time-series were calculated based on the preprocessed BOLD: framewise displacement (FD), DVARS and three region-wise global signals. FD was computed using two formulations following Power (absolute sum of relative motions)[13] and Jenkinson (relative root mean square displacement between affines)[11]. FD and DVARS are calculated for each functional run, both using their implementations in Nipype[13]. The three global signals are extracted within the CSF, the WM, and the whole-brain masks. Additionally, a set of physiological regressors were extracted to allow for component-based noise correction (CompCor)[14]. Principal components are estimated after high-pass filtering the preprocessed BOLD time-series (using a discrete cosine filter with 128s cut-off) for the two CompCor variants: temporal (tCompCor) and anatomical (aCompCor). tCompCor components are then calculated from the top 2% variable voxels within the brain mask. For aCompCor, three probabilistic masks (CSF, WM and combined CSF+WM) are generated in anatomical space. The implementation differs from that of Behzadi et al.[14] in that instead of eroding the masks by 2 pixels on BOLD space, the aCompCor masks are subtracted a mask of pixels that likely contain a volume fraction of GM. This mask is obtained by dilating a GM mask extracted from the FreeSurfer's aseg segmentation, and it ensures components are not extracted from voxels containing a minimal fraction of GM. Finally, these masks are resampled into BOLD space and binarized by thresholding at 0.99 (as in the original implementation). Components are also calculated separately within the WM and CSF masks. For each CompCor decomposition, the k components with the largest singular values are retained, such that the retained components' time series are sufficient to explain 50 percent of variance across the nuisance mask (CSF, WM, combined, or temporal). The remaining components are dropped from consideration. The head-motion estimates calculated in the correction step were also placed within the corresponding confounds file. The confound time series derived from head motion estimates and global signals were expanded with the inclusion of temporal derivatives and quadratic terms for each (Satterthwaite et al. 2013). Frames that exceeded a threshold of 0.5 mm FD or 1.5 standardised DVARS were annotated as motion outliers. All resamplings can be performed with a single interpolation step by composing all the pertinent transformations (i.e. head-motion transform matrices, susceptibility distortion correction when available, and co-registrations to anatomical and output spaces). Gridded (volumetric) resamplings were performed using antsApplyTransforms (ANTs), configured with Lanczos interpolation to minimize the smoothing effects of other kernels (Lanczos 1964). Non-gridded (surface) resamplings were performed using mri_vol2surf (FreeSurfer).

The output of the fMRIPrep pre-processing pipeline was subsequently input into the xcpEngine post-processing pipeline[15]. The xcpEngine post-processing pipeline is a self-contained software that allows the rapid and reproducible implementation of tools necessary for calculating functional connectivity, while also allowing for benchmarking pipeline performance using a wide array of benchmarking pipelines[16]. xcpEngine is built to use the output of the fMRIPrep pipeline as an input, therefore, many of the pipeline steps use metrics explicitly calculated by fMRIPrep. Briefly, the steps implemented in the xcpEngine pipeline for each subject, were as follows. First, regressors for artifactual signals were calculated from the 4D time series of each subject using the confound2 module. The regressed parameters calculated by this module included motion realignment parameters (3 rotational and 3 translational) necessary for realigning each volume in the time series to a reference volume; the mean white matter and cerebrospinal fluid time series over all voxels[17], with tissue segmentations determined by the fMRIPrep; the mean time series signal across the whole brain[13]; the temporal derivative of motion parameters, which encodes the relative displacement of the brain from one volume of the timeseries to the next[17]; and finally, the second power of each of the previously mentioned regressors was also included, to account for potential noise that is proportional to higher powers of motion and nuisance regressors (a total of 36 regression parameters). After estimating the regressors, demeaning and detrending, followed by temporal filtering was carried out in both the BOLD timeseries and the regressors, using the regress module. The timeseries and regressors were detrended using a 2nd order polynomial. A first order forward-backward bandpass Butterworth filter, with passband 0.01-0.10Hz was implemented, allowing both high frequency noise, and very-low-frequency drift to be eliminated. The filtered regressors were fitted to the filtered BOLD timeseries data using multiple linear regression. Any variance in the BOLD timeseries explained by the regressors was discarded from the timeseries, whereas the unexplained variance was left as the final filtered timeseries.

**Identification of the anterior commissure and shape analysis**

A deterministic fiber tracking algorithm[18] was used with augmented tracking strategies[19] to improve reproducibility. The anisotropy threshold was randomly selected. The angular threshold was randomly selected from 15 degrees to 90 degrees. The step size was randomly selected from 0.5 voxel to 1.5 voxels. The fiber trajectories were smoothed by averaging the propagation direction with a percentage of the previous direction. The percentage was randomly selected from 0% to 95%. We evaluated the effects of omitting this smoothing procedure. Tracks with length shorter than 30 or longer than 200 mm were discarded. A total of 100,000 seeds were placed. Shape analysis[19] was conducted to derive shape metrics for tractography using a fixed Otsu threshold of 0.5. We evaluated the effects of AutoTrack tolerance, defined as the Hausdorff distance of streamlines from a coregistered anterior commissure template from the HCP842 tractography atlas[20], on shape metrics. parameter_id=cba3Fb803Fcb803FbF041b4843A08601eca.

1492(199706/08)10:4/5<171::AID-NBM453>3.0.CO;2-L

**Supplementary Table 1 - Subject demographics separated by IPC group**

### Subject Demographics

| Characteristic | Healthy Controls | High-IPC | Low-IPC | BiTLE |
|---|---|---|---|---|
| **Total Subjects** | 26 | 36 | 19 | 8 |
| **Age** | 30±10 | 39±11 | 43±10 | 35±13 |
| **Female** | 12 | 21 | 10 | 4 |
| **Disease Duration (years)** | - | 17±16 | 15±13 | 13±8 |
| **Lesional MRI** | - | 22 | 13 | 4 |
| **Disease Laterality** | | | | |
| *Left* | - | 22 | 12 | - |
| *Right* | - | 14 | 7 | - |
| *Bilateral* | - | - | - | 8 |
| **Surgical Outcomes (24 months)** | | | | |
| *Engel IA* | - | 6 | 1 | 0 |
| *Engel IB-IV* | - | 12 | 8 | 2 |
| *Unavailable* | - | 18 | 10 | 6 |
| **Acquisition Protocol** | | | | |
| *Version A* | 7 | 7 | 3 | 0 |
| *Version B* | 19 | 29 | 16 | 8 |

**Supplementary Table 2 – Emory subject demographics**

### Subject Demographics

| Characteristic | Healthy Controls | TLE |
|---|---|---|
| **Total Subjects** | 10 | 28 |
| **Age** | 36±18 | 38±9 |
| **Female** | 12 | 18 |
| **Disease Duration (years)** | - | 20±15 |
| **Lesional MRI** | - | 12 |
| **Disease Laterality** | | |
| *Left* | - | 13 |
| *Right* | - | 16 |

**Supplementary Table 3**: Percent overlap between the Harvard-Oxford cortical and subcortical atlas brain regions and the functional connectivity mask of the right piriform > right hippocampus in the HCP cohort

| Brain Region | Percent Overlap (%) |
|---|---|
| Insular Cortex | 7.8543 |
| Frontal Pole | 4.6544 |
| Left Putamen | 4.4158 |
| Right Putamen | 4.2463 |
| Inferior Frontal Gyrus, pars opercularis | 3.8043 |
| Cingulate Gyrus, anterior division | 3.7624 |
| Supramarginal Gyrus, anterior division | 3.4184 |
| Precentral Gyrus | 3.384 |
| Central Opercular Cortex | 2.7914 |
| Frontal Operculum Cortex | 2.6342 |
| Superior Frontal Gyrus | 2.2944 |
| Inferior Frontal Gyrus, pars triangularis | 2.1625 |
| Middle Frontal Gyrus | 2.0385 |
| Paracingulate Gyrus | 1.955 |
| Left Caudate | 1.6993 |
| Juxtapositional Lobule Cortex (formerly Supplementary Motor Cortex) | 1.4566 |
| Supramarginal Gyrus, posterior division | 1.4149 |
| Frontal Orbital Cortex | 1.3031 |
| Right Thalamus | 1.2961 |
| Right Caudate | 0.9811 |
| Parietal Operculum Cortex | 0.7208 |
| Left Amygdala | 0.7032 |
| Temporal Pole | 0.6881 |
| Postcentral Gyrus | 0.5408 |
| Planum Polare | 0.495 |
| Right Amygdala | 0.4888 |
| Planum Temporale | 0.4016 |
| Superior Temporal Gyrus, posterior division | 0.2636 |
| Left Lateral Ventricle | 0.2482 |
| Right Lateral Ventricle | 0.2224 |
| Angular Gyrus | 0.1539 |
| Parahippocampal Gyrus, anterior division | 0.0841 |
| Right Pallidum | 0.0785 |
| Heschl's Gyrus (includes H1 and H2) | 0.0749 |
| Left Pallidum | 0.0625 |

| | |
|---|---|
| Left Accumbens | 0.0371 |
| Left Thalamus | 0.015 |
| Superior Temporal Gyrus, anterior division | 0.011 |
| Superior Parietal Lobule | 0.0056 |
| Cingulate Gyrus, posterior division | 0.003 |
| Right Accumbens | 0.0022 |
| Subcallosal Cortex | 0.0004 |
| Left Hippocampus | 0.0004 |
| Right Hippocampus | 0.0001 |

**Supplementary Table 4**: Percent overlap between the Harvard-Oxford cortical and subcortical atlas brain regions and the functional connectivity mask of the left piriform > left hippocampus in the HCP cohort

| Brain Region | Percent Overlap (%) |
|---|---:|
| Insular Cortex | 6.5987 |
| Frontal Pole | 6.4391 |
| Central Opercular Cortex | 3.5589 |
| Supramarginal Gyrus, anterior division | 3.3748 |
| Cingulate Gyrus, anterior division | 3.3602 |
| Precentral Gyrus | 3.1682 |
| Inferior Frontal Gyrus, pars opercularis | 3.0146 |
| Left Putamen | 2.993 |
| Right Putamen | 2.8956 |
| Supramarginal Gyrus, posterior division | 2.4398 |
| Frontal Operculum Cortex | 2.4316 |
| Juxtapositional Lobule Cortex (formerly Supplementary Motor Cortex) | 2.3434 |
| Frontal Orbital Cortex | 2.3298 |
| Parietal Operculum Cortex | 1.8848 |
| Inferior Frontal Gyrus, pars triangularis | 1.6814 |
| Planum Polare | 1.6399 |
| Paracingulate Gyrus | 1.6277 |
| Superior Frontal Gyrus | 1.5649 |
| Left Caudate | 1.4155 |
| Postcentral Gyrus | 1.3705 |
| Right Amygdala | 1.1291 |
| Temporal Pole | 1.0889 |
| Planum Temporale | 1.0057 |
| Middle Frontal Gyrus | 1.0044 |
| Angular Gyrus | 0.8194 |
| Right Caudate | 0.4612 |
| Superior Temporal Gyrus, posterior division | 0.4072 |
| Heschl's Gyrus (includes H1 and H2) | 0.3968 |
| Left Amygdala | 0.3562 |
| Left Lateral Ventricle | 0.2795 |
| Parahippocampal Gyrus, anterior division | 0.1673 |
| Left Thalamus | 0.0857 |
| Superior Temporal Gyrus, anterior division | 0.077 |
| Right Pallidum | 0.0543 |
| Subcallosal Cortex | 0.0527 |
| Right Lateral Ventricle | 0.0513 |

| | |
|---|---|
| Superior Parietal Lobule | 0.0448 |
| Left Pallidum | 0.044 |
| Cingulate Gyrus, posterior division | 0.0419 |
| Left Accumbens | 0.0418 |
| Precuneous Cortex | 0.0417 |
| Lateral Occipital Cortex, superior division | 0.0284 |
| Right Hippocampus | 0.0168 |
| Right Thalamus | 0.0106 |
| Right Accumbens | 0.0041 |
| Middle Temporal Gyrus, temporooccipital part | 0.0024 |
| Middle Temporal Gyrus, posterior division | 0.0008 |
| Left Hippocampus | 0.0001 |

**Supplementary Table 5**: Percent overlap between the Harvard-Oxford cortical and subcortical atlas brain regions and the functional connectivity mask of the right hippocampus > right piriform in the HCP cohort

| Brain Region | Percent Overlap (%) |
|---|---|
| Frontal Pole | 5.8667 |
| Lateral Occipital Cortex, superior division | 5.067 |
| Precuneous Cortex | 4.3699 |
| Temporal Pole | 3.4613 |
| Postcentral Gyrus | 2.5628 |
| Middle Temporal Gyrus, posterior division | 2.4415 |
| Lateral Occipital Cortex, inferior division | 2.3071 |
| Cingulate Gyrus, posterior division | 2.2772 |
| Precentral Gyrus | 2.1314 |
| Paracingulate Gyrus | 2.0283 |
| Lingual Gyrus | 1.9264 |
| Superior Frontal Gyrus | 1.7416 |
| Superior Temporal Gyrus, posterior division | 1.5701 |
| Temporal Fusiform Cortex, posterior division | 1.5198 |
| Angular Gyrus | 1.3798 |
| Middle Frontal Gyrus | 1.3734 |
| Frontal Medial Cortex | 1.3156 |
| Inferior Temporal Gyrus, posterior division | 1.2233 |
| Parahippocampal Gyrus, anterior division | 1.1759 |
| Frontal Orbital Cortex | 1.1692 |
| Cuneal Cortex | 1.1588 |
| Occipital Pole | 1.1536 |
| Subcallosal Cortex | 1.1288 |
| Cingulate Gyrus, anterior division | 1.0698 |
| Left Hippocampus | 1.0557 |
| Right Hippocampus | 1.0523 |
| Temporal Occipital Fusiform Cortex | 1.0056 |
| Middle Temporal Gyrus, temporooccipital part | 0.923 |
| Middle Temporal Gyrus, anterior division | 0.9163 |
| Inferior Temporal Gyrus, temporooccipital part | 0.8852 |
| Parahippocampal Gyrus, posterior division | 0.8273 |
| Occipital Fusiform Gyrus | 0.8211 |
| Superior Parietal Lobule | 0.5774 |
| Superior Temporal Gyrus, anterior division | 0.5547 |
| Intracalcarine Cortex | 0.5294 |

| | |
|---|---|
| Supracalcarine Cortex | 0.4914 |
| Supramarginal Gyrus, posterior division | 0.4011 |
| Temporal Fusiform Cortex, anterior division | 0.3817 |
| Inferior Temporal Gyrus, anterior division | 0.3494 |
| Left Amygdala | 0.3277 |
| Left Thalamus | 0.2821 |
| Right Amygdala | 0.2817 |
| Supramarginal Gyrus, anterior division | 0.1821 |
| Planum Temporale | 0.1194 |
| Right Thalamus | 0.1172 |
| Right Accumbens | 0.1038 |
| Left Accumbens | 0.085 |
| Juxtapositional Lobule Cortex (formerly Supplementary Motor Cortex) | 0.0741 |
| Left Lateral Ventricle | 0.0709 |
| Right Lateral Ventricle | 0.0681 |
| Brain-Stem | 0.0674 |
| Planum Polare | 0.0473 |
| Heschl's Gyrus (includes H1 and H2) | 0.0243 |
| Insular Cortex | 0.0218 |
| Right Caudate | 0.0163 |
| Left Caudate | 0.0137 |
| Central Opercular Cortex | 0.0117 |
| Inferior Frontal Gyrus, pars triangularis | 0.0093 |
| Left Putamen | 0.0026 |
| Parietal Operculum Cortex | 0.0025 |
| Right Putamen | 0.0022 |
| Inferior Frontal Gyrus, pars opercularis | 0.0019 |
| Frontal Operculum Cortex | 0.0014 |
| Left Pallidum | 0.0007 |
| Right Pallidum | 0.0003 |

**Supplementary Table 6:** Percent overlap between the Harvard-Oxford cortical and subcortical atlas brain regions and the functional connectivity mask of the left hippocampus > left piriform in the HCP cohort

| **Brain Region** | **Percent Overlap (%)** |
|---|---|
| Frontal Pole | 6.9034 |
| Precuneous Cortex | 4.9653 |
| Lateral Occipital Cortex, superior division | 4.913 |
| Temporal Pole | 3.2733 |
| Middle Temporal Gyrus, posterior division | 2.6428 |
| Cingulate Gyrus, posterior division | 2.6105 |
| Lingual Gyrus | 2.5175 |
| Paracingulate Gyrus | 2.3388 |
| Superior Frontal Gyrus | 2.235 |
| Middle Frontal Gyrus | 1.8246 |
| Precentral Gyrus | 1.8023 |
| Temporal Fusiform Cortex, posterior division | 1.5712 |
| Frontal Medial Cortex | 1.5703 |
| Postcentral Gyrus | 1.4924 |
| Inferior Temporal Gyrus, posterior division | 1.4175 |
| Subcallosal Cortex | 1.3139 |
| Superior Temporal Gyrus, posterior division | 1.2665 |
| Left Hippocampus | 1.2653 |
| Right Hippocampus | 1.2514 |
| Angular Gyrus | 1.2406 |
| Lateral Occipital Cortex, inferior division | 1.1787 |
| Middle Temporal Gyrus, anterior division | 1.114 |
| Cingulate Gyrus, anterior division | 1.1027 |
| Parahippocampal Gyrus, anterior division | 1.0815 |
| Parahippocampal Gyrus, posterior division | 0.9918 |
| Frontal Orbital Cortex | 0.9778 |
| Temporal Occipital Fusiform Cortex | 0.9476 |
| Cuneal Cortex | 0.8517 |
| Occipital Fusiform Gyrus | 0.7815 |
| Middle Temporal Gyrus, temporooccipital part | 0.6886 |
| Intracalcarine Cortex | 0.6754 |
| Occipital Pole | 0.6145 |
| Superior Temporal Gyrus, anterior division | 0.5983 |
| Inferior Temporal Gyrus, temporooccipital part | 0.5837 |
| Supracalcarine Cortex | 0.5521 |

| | |
|---|---|
| Inferior Temporal Gyrus, anterior division | 0.4229 |
| Left Amygdala | 0.3376 |
| Supramarginal Gyrus, posterior division | 0.2961 |
| Temporal Fusiform Cortex, anterior division | 0.2725 |
| Right Thalamus | 0.2331 |
| Superior Parietal Lobule | 0.1786 |
| Left Thalamus | 0.1712 |
| Right Amygdala | 0.1613 |
| Right Accumbens | 0.1191 |
| Right Lateral Ventricle | 0.0966 |
| Left Lateral Ventricle | 0.0711 |
| Supramarginal Gyrus, anterior division | 0.0659 |
| Left Accumbens | 0.0643 |
| Juxtapositional Lobule Cortex (formerly Supplementary Motor Cortex) | 0.0518 |
| Brain-Stem | 0.0491 |
| Inferior Frontal Gyrus, pars triangularis | 0.0487 |
| Planum Temporale | 0.0438 |
| Inferior Frontal Gyrus, pars opercularis | 0.0222 |
| Planum Polare | 0.021 |
| Right Caudate | 0.0162 |
| Heschl's Gyrus (includes H1 and H2) | 0.0087 |
| Left Caudate | 0.0035 |
| Parietal Operculum Cortex | 0.0019 |
| Right Putamen | 0.0018 |
| Left Putamen | 0.0011 |
| Frontal Operculum Cortex | 0.0009 |
| Central Opercular Cortex | 0.0005 |
| Insular Cortex | 0.0003 |
| Right Pallidum | 0.0003 |
| Left Pallidum | 0.0002 |

**Supplementary Table 7:** Percent overlap between the Harvard-Oxford cortical and subcortical atlas brain regions and the functional connectivity mask of the right piriform > right hippocampus in the Penn cohort

| Brain Region | Percent Overlap (%) |
|---|---|
| Insular Cortex | 17.0787 |
| Central Opercular Cortex | 8.3394 |
| Right Putamen | 7.5962 |
| Inferior Frontal Gyrus, pars opercularis | 6.2744 |
| Right Amygdala | 5.8245 |
| Precentral Gyrus | 4.3055 |
| Temporal Pole | 3.4061 |
| Frontal Operculum Cortex | 2.9096 |
| Frontal Orbital Cortex | 1.6387 |
| Parahippocampal Gyrus, anterior division | 1.0191 |
| Planum Polare | 0.7007 |
| Right Caudate | 0.3421 |
| Inferior Frontal Gyrus, pars triangularis | 0.1968 |
| Right Pallidum | 0.126 |
| Middle Frontal Gyrus | 0.1181 |
| Supramarginal Gyrus, anterior division | 0.1118 |
| Parietal Operculum Cortex | 0.0561 |
| Heschl's Gyrus (includes H1 and H2) | 0.0386 |
| Planum Temporale | 0.0281 |
| Right Hippocampus | 0.026 |
| Right Lateral Ventricle | 0.0256 |
| Right Accumbens | 0.0221 |
| Superior Temporal Gyrus, posterior division | 0.0171 |
| Left Putamen | 0.0135 |
| Postcentral Gyrus | 0.0055 |
| Superior Temporal Gyrus, anterior division | 0.0047 |
| Subcallosal Cortex | 0.0035 |
| Supramarginal Gyrus, posterior division | 0.0027 |

**Supplementary Table 8:** Percent overlap between the Harvard-Oxford cortical and subcortical atlas brain regions and the functional connectivity mask of the left piriform > left hippocampus in the Penn cohort

| Brain Region | Percent Overlap (%) |
|---|---|
| Insular Cortex | 15.0413 |
| Central Opercular Cortex | 7.2421 |
| Left Putamen | 6.7792 |
| Right Putamen | 5.7215 |
| Frontal Operculum Cortex | 3.5295 |
| Inferior Frontal Gyrus, pars opercularis | 3.3828 |
| Precentral Gyrus | 3.2931 |
| Left Amygdala | 3.1154 |
| Right Amygdala | 3.0557 |
| Temporal Pole | 2.4953 |
| Frontal Orbital Cortex | 2.1215 |
| Planum Polare | 1.3186 |
| Parahippocampal Gyrus, anterior division | 1.1292 |
| Right Thalamus | 0.9025 |
| Heschl's Gyrus (includes H1 and H2) | 0.6448 |
| Parietal Operculum Cortex | 0.6384 |
| Left Thalamus | 0.5718 |
| Supramarginal Gyrus, anterior division | 0.5687 |
| Planum Temporale | 0.2738 |
| Inferior Frontal Gyrus, pars triangularis | 0.146 |
| Supramarginal Gyrus, posterior division | 0.1391 |
| Left Pallidum | 0.1093 |
| Left Hippocampus | 0.1008 |
| Right Pallidum | 0.0924 |
| Superior Temporal Gyrus, posterior division | 0.0772 |
| Postcentral Gyrus | 0.0526 |
| Superior Temporal Gyrus, anterior division | 0.0212 |
| Angular Gyrus | 0.0174 |
| Right Hippocampus | 0.0147 |
| Left Accumbens | 0.0096 |
| Right Caudate | 0.0078 |
| Middle Frontal Gyrus | 0.0075 |
| Middle Temporal Gyrus, temporooccipital part | 0.0038 |
| Middle Temporal Gyrus, posterior division | 0.0027 |

| | |
|---|---|
| Right Lateral Ventricle | 0.0011 |
| Right Accumbens | 0.0009 |
| Subcallosal Cortex | 0.0008 |
| Left Lateral Ventricle | 0.0004 |
| Left Caudate | 0.0003 |

**Supplementary Table 9**: Percent overlap between the Harvard-Oxford cortical and subcortical atlas brain regions and the functional connectivity mask of the right hippocampus > right piriform in the Penn cohort

| Brain Region | Percent Overlap (%) |
|---|---|
| Precuneous Cortex | 10.2561 |
| Frontal Pole | 9.8311 |
| Cingulate Gyrus, posterior division | 9.5788 |
| Frontal Medial Cortex | 5.5106 |
| Right Hippocampus | 5.1824 |
| Paracingulate Gyrus | 5.0813 |
| Left Hippocampus | 4.926 |
| Parahippocampal Gyrus, anterior division | 3.5735 |
| Parahippocampal Gyrus, posterior division | 3.5094 |
| Temporal Fusiform Cortex, posterior division | 2.2633 |
| Subcallosal Cortex | 2.2585 |
| Cingulate Gyrus, anterior division | 1.7876 |
| Lingual Gyrus | 1.5501 |
| Left Thalamus | 1.1121 |
| Right Thalamus | 1.0733 |
| Supracalcarine Cortex | 0.8913 |
| Intracalcarine Cortex | 0.501 |
| Right Amygdala | 0.4076 |
| Cuneal Cortex | 0.3592 |
| Temporal Fusiform Cortex, anterior division | 0.3321 |
| Frontal Orbital Cortex | 0.3247 |
| Left Amygdala | 0.2526 |
| Right Lateral Ventricle | 0.2314 |
| Inferior Temporal Gyrus, posterior division | 0.2213 |
| Temporal Pole | 0.2031 |
| Superior Frontal Gyrus | 0.1748 |
| Left Lateral Ventricle | 0.1685 |
| Temporal Occipital Fusiform Cortex | 0.1615 |
| Brain-Stem | 0.1086 |
| Inferior Temporal Gyrus, anterior division | 0.0175 |
| Precentral Gyrus | 0.0119 |
| Right Putamen | 0.0017 |
| Postcentral Gyrus | 0.0006 |
| Left Putamen | 0.0005 |
| Superior Parietal Lobule | 0.0002 |

| | |
|---|---|
| Lateral Occipital Cortex, superior division | 0.0002 |
| Inferior Temporal Gyrus, temporooccipital part | 0.0001 |
| Left Caudate | 0 |
| Left Accumbens | 0 |
| Right Pallidum | 0 |

**Supplementary Table 10**: Percent overlap between the Harvard-Oxford cortical and subcortical atlas brain regions and the functional connectivity mask of the left hippocampus > left piriform in the Penn cohort

| Brain Region | Percent Overlap (%) |
|---|---|
| Frontal Pole | 11.4841 |
| Precuneous Cortex | 6.4516 |
| Cingulate Gyrus, posterior division | 6.3671 |
| Paracingulate Gyrus | 5.9304 |
| Middle Temporal Gyrus, posterior division | 4.0883 |
| Frontal Medial Cortex | 4.0206 |
| Left Hippocampus | 3.2896 |
| Right Hippocampus | 2.9564 |
| Inferior Temporal Gyrus, posterior division | 2.8311 |
| Superior Frontal Gyrus | 2.6301 |
| Parahippocampal Gyrus, anterior division | 2.3407 |
| Middle Temporal Gyrus, anterior division | 2.1357 |
| Cingulate Gyrus, anterior division | 2.0534 |
| Temporal Fusiform Cortex, posterior division | 1.7009 |
| Subcallosal Cortex | 1.4637 |
| Parahippocampal Gyrus, posterior division | 1.3419 |
| Inferior Temporal Gyrus, anterior division | 1.1093 |
| Left Thalamus | 0.8191 |
| Middle Frontal Gyrus | 0.6134 |
| Temporal Pole | 0.5563 |
| Superior Temporal Gyrus, posterior division | 0.5495 |
| Temporal Fusiform Cortex, anterior division | 0.5467 |
| Middle Temporal Gyrus, temporooccipital part | 0.3303 |
| Lingual Gyrus | 0.3172 |
| Frontal Orbital Cortex | 0.3165 |
| Superior Temporal Gyrus, anterior division | 0.2778 |
| Supracalcarine Cortex | 0.2746 |
| Right Thalamus | 0.2667 |
| Left Amygdala | 0.2149 |
| Cuneal Cortex | 0.184 |
| Intracalcarine Cortex | 0.1743 |
| Left Lateral Ventricle | 0.1321 |
| Inferior Temporal Gyrus, temporooccipital part | 0.1126 |
| Right Amygdala | 0.0682 |
| Right Lateral Ventricle | 0.0517 |

| | |
|---|---|
| Precentral Gyrus | 0.0104 |
| Brain-Stem | 0.0083 |
| Temporal Occipital Fusiform Cortex | 0.0079 |
| Planum Temporale | 0.0023 |
| Left Putamen | 0.002 |
| Postcentral Gyrus | 0.0012 |
| Lateral Occipital Cortex, superior division | 0.0011 |
| Superior Parietal Lobule | 0.0009 |
| Angular Gyrus | 0.0007 |
| Planum Polare | 0.0006 |
| Supramarginal Gyrus, posterior division | 0.0003 |
| Heschl's Gyrus (includes H1 and H2) | 0.0002 |
| Left Accumbens | 0.0002 |
| Right Putamen | 0.0002 |
| Lateral Occipital Cortex, inferior division | 0.0001 |
| Left Pallidum | 0.0001 |
| Left Caudate | 0 |

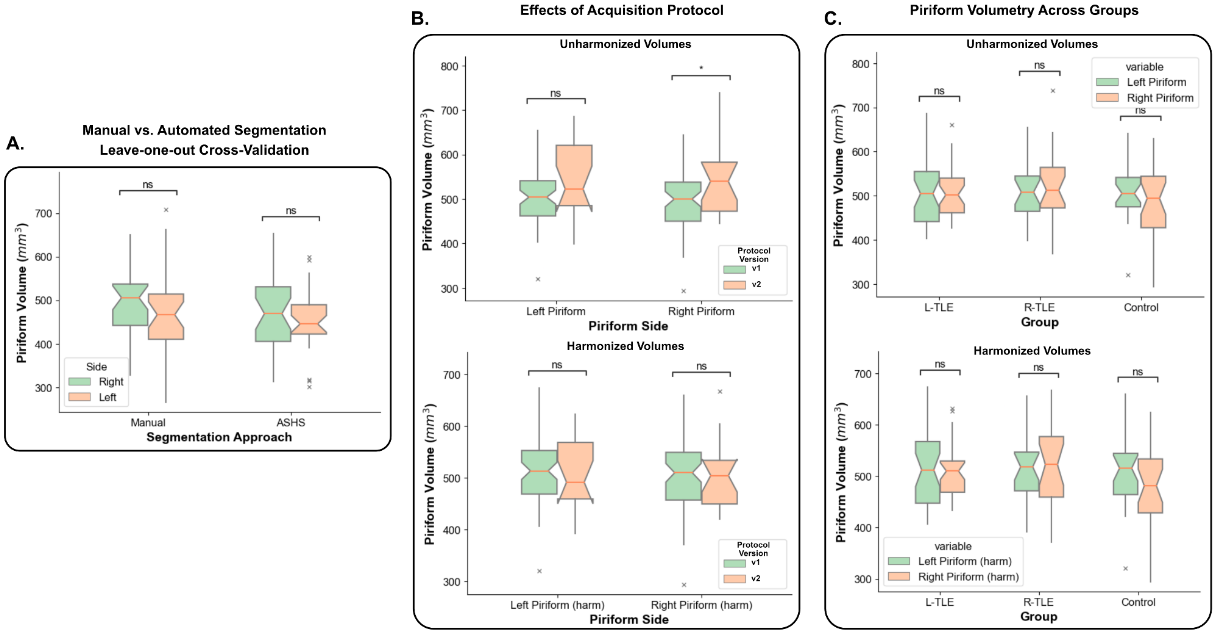


**Supplementary Figure 1 - Piriform Volumetry: Panel A.** shows the results of the leave-one-out cross-validation in the ASHS model trained to perform piriform segmentation. **Panel B.** shows the effects of the acquisition protocols used in the resulting piriform volumetry, and the removal of those effects after harmonization of the data. **Panel C.** shows the left and right piriform volumetry for control, L-TLE and R-TLE subjects before and after harmonization. We report the harmonized values in the main text. n.s. - not significant. *p<0.05, **p<0.01, ***p<0.001, ****p<0.0001. *p*-values are Bonferroni corrected where appropriate.

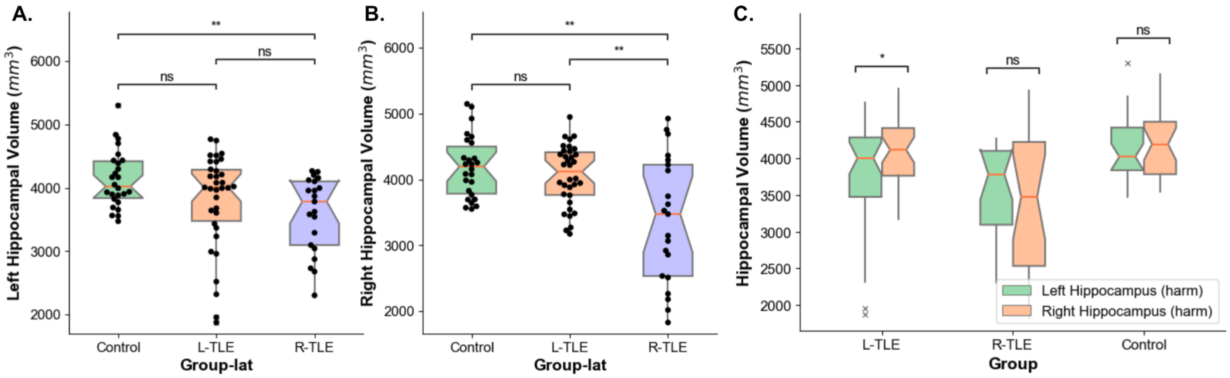


**Supplementary Figure 2 - Hippocampal volumetry:** Volumetry computed from an ASHS segmentation of the (**A.**) left and (**B.**) right hippocampus for control, L-TLE and R-TLE subjects. **Panel C.** shows the volumes grouped by subject group. All volumes reported here are harmonized between the two acquisition protocols. n.s. - not significant. *p<0.05, **p<0.01, ***p<0.001, ****p<0.0001. *p*-values are Bonferroni corrected where appropriate.

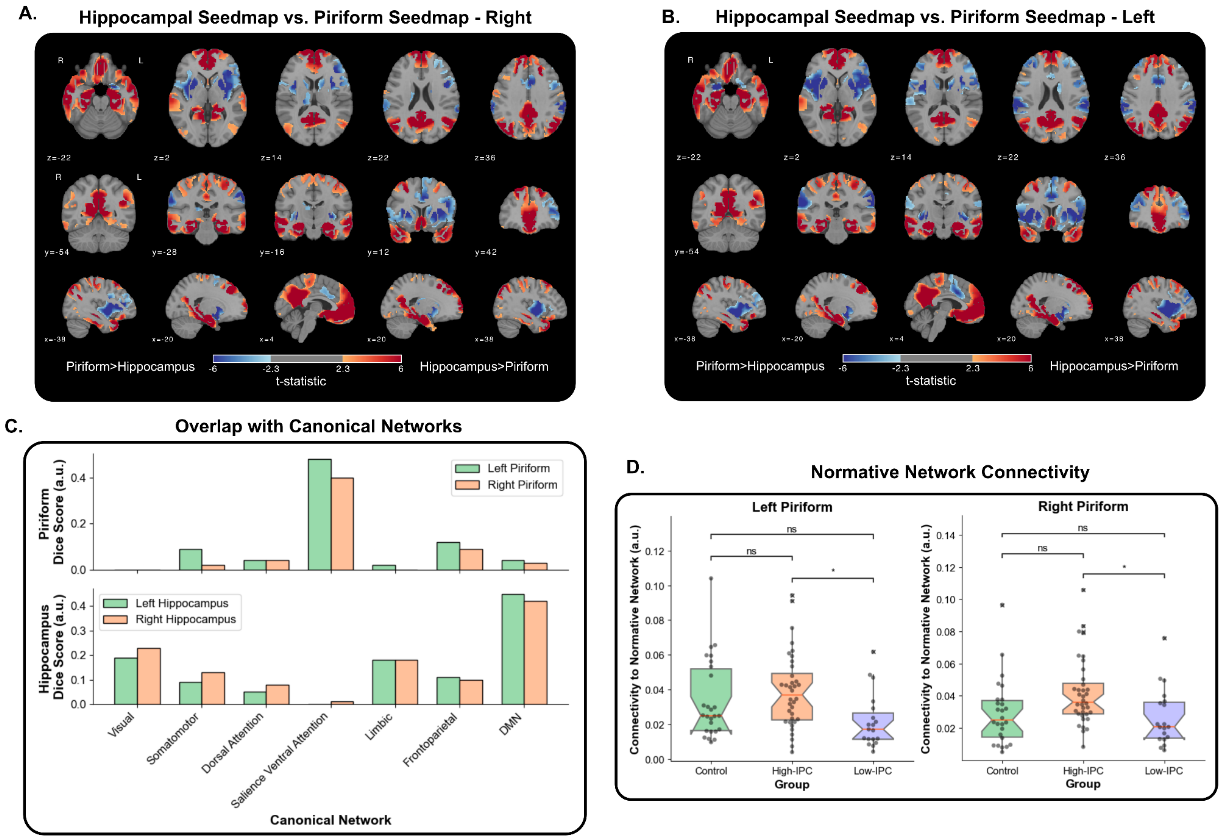


**Supplementary Figure 3 - Normative piriform connectivity in the human connectome project:** We replicated the functional connectivity comparison between the piriform and the hippocampus presented in **Figure 2** of the main text, but in a subset of 100 human connectome project (HCP) subjects. **Panels A. and B.** demonstrate the voxelwise comparison between the piriform and the hippocampal seedmaps in healthy control subjects for both right (**A.**) and left (**B.**) hippocampus and piriform. Shown voxels survived threshold-free cluster enhancement significance testing ($p<0.05$), and the colormap represents the corresponding t-statistic of each voxel. **Panel C.** shows the Dice overlap score between the normative piriform mask (top) and the normative hippocampal mask (bottom) across the Yeo-Krienen canonical networks. **Panel E.** shows an analogous result to **Figure 4E** of the main text**,** but for the normative mask created from HCP subjects. n.s. - not significant. *$p<0.05$, **$p<0.01$, ***$p<0.001$, ****$p<0.0001$. *p*-values are Bonferroni corrected where appropriate.

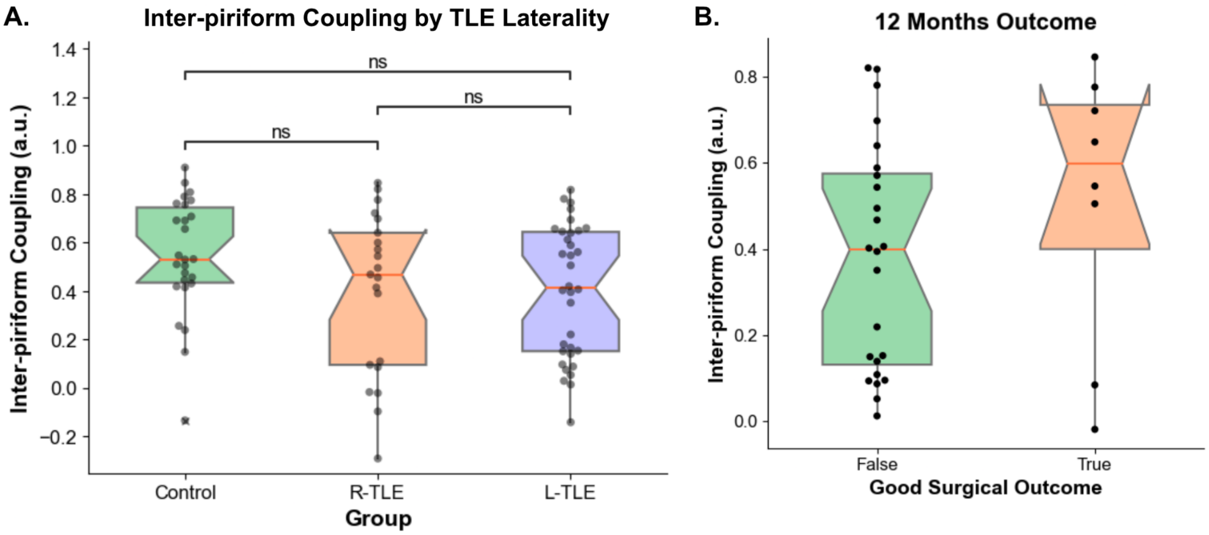


**Supplementary Figure 4 - Inter-piriform coupling by TLE laterality and 12 months outcomes: Panel A.** shows the IPC comparison between control, R-TLE and L-TLE subjects. **Panel B.** shows the IPC between patients with good (Engel IA) and poor (Engel IB-IV) 12 month surgical outcomes. n.s. - not significant. *p<0.05, **p<0.01, ***p<0.001, ****p<0.0001. *p*-values are Bonferroni corrected where appropriate.

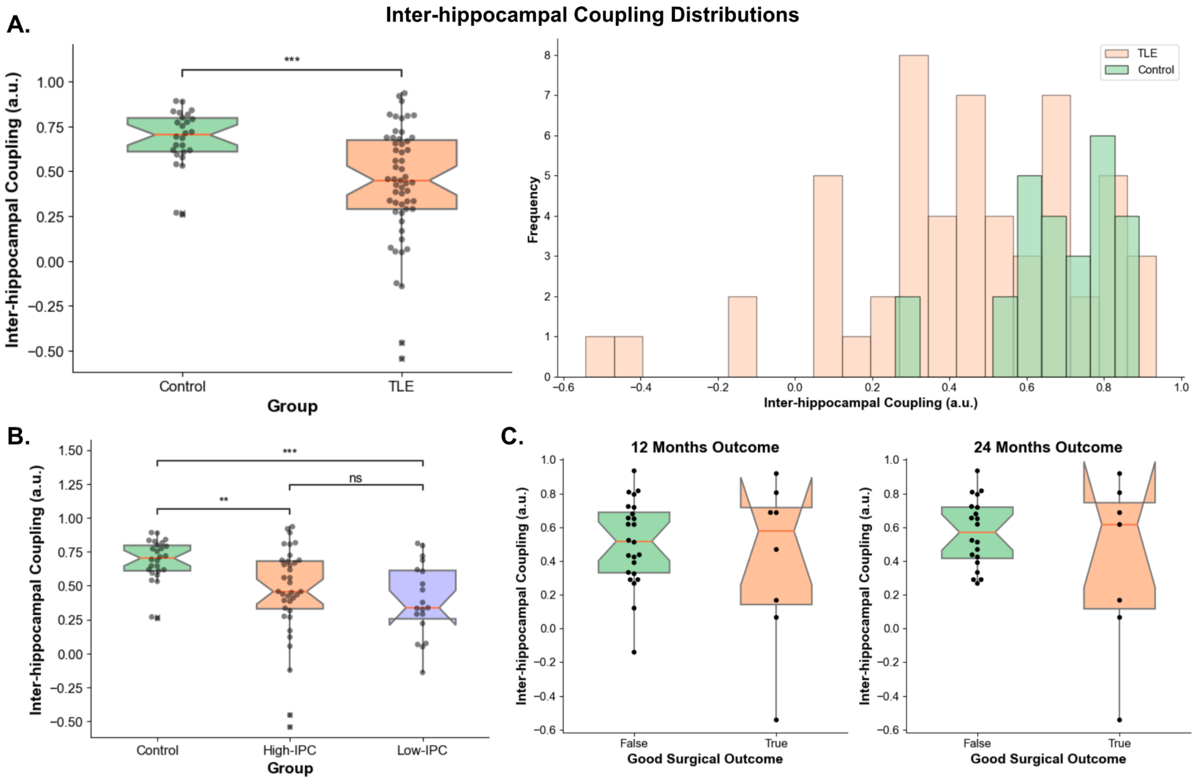


**Supplementary Figure 5 - Inter-hippocampal coupling: Panel A.** shows the inter-hippocampal coupling between control and TLE subjects. **Panel B.** shows the inter-hippocampal coupling between the control, high and low-IPC groups. **Panel C.** shows the inter-hippocampal coupling between patients with good (Engel IA) and poor (Engel IB-IV) 12 month (left) and 24 month (right) surgical outcomes. n.s. - not significant. *p<0.05, **p<0.01, ***p<0.001, ****p<0.0001. *p*-values are Bonferroni corrected where appropriate.

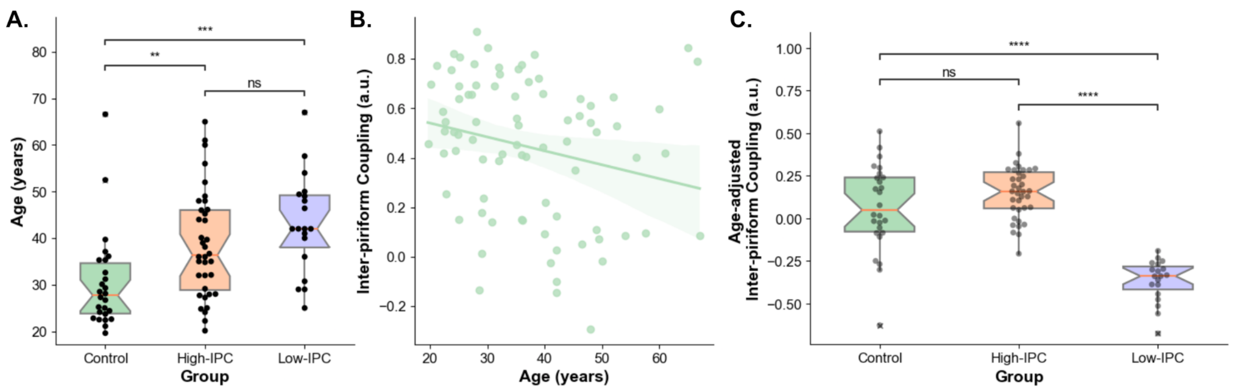


**Supplementary Figure 6 - Effect of age on inter-piriform coupling: Panel A.** shows the age for subjects in the control, high and low-IPC groups. **Panel B.** shows the scatterplot between IPC and age (Spearman's $r$ = -0.28, $p$=0.011). **Panel C.** shows the IPC values for the same groups but after age was linearly regressed out. n.s. - not significant. *p<0.05, **p<0.01, ***p<0.001, ****p<0.0001. $p$-values are Bonferroni corrected.

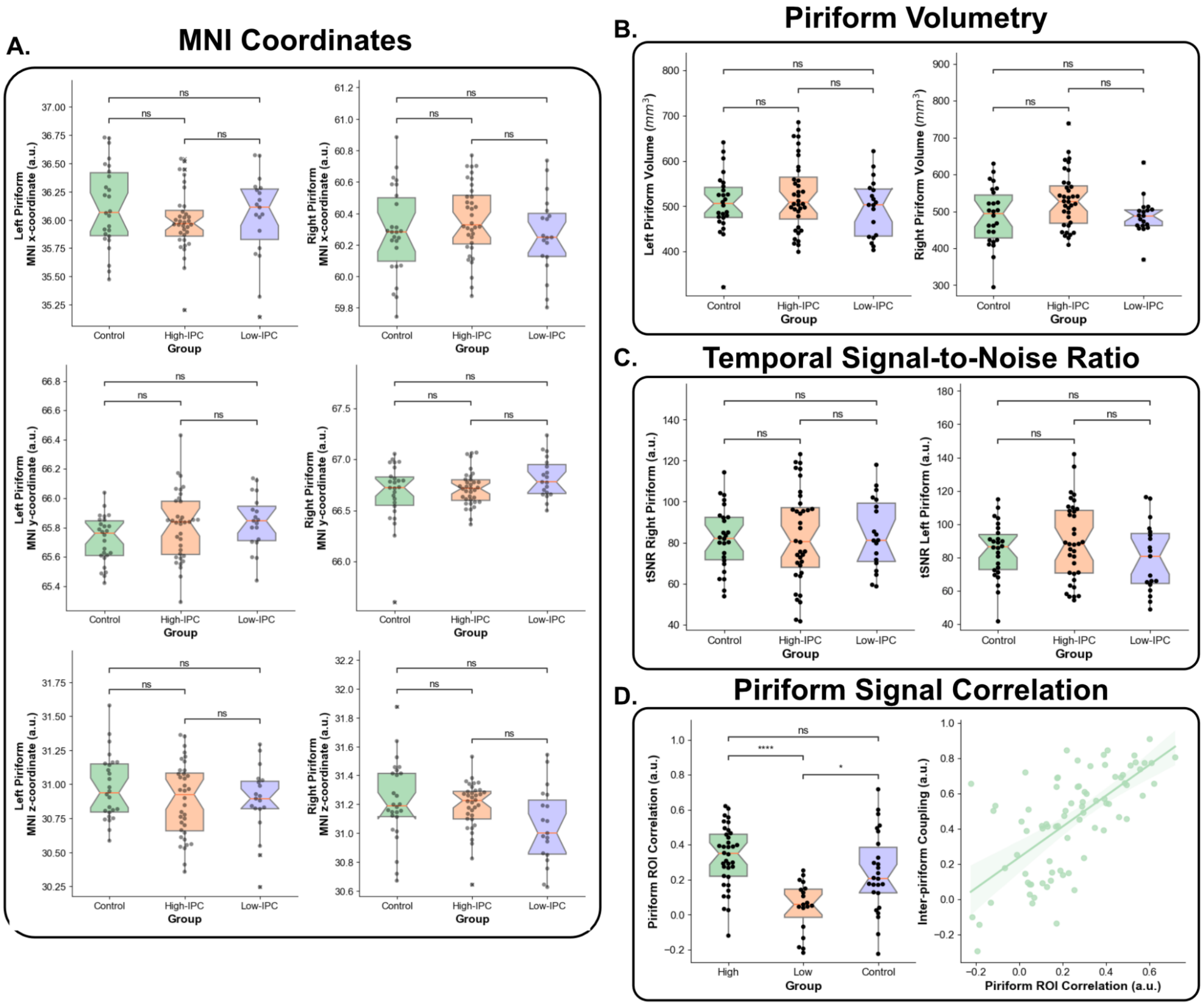


**Supplementary Figure 7 - Assessment of potential structural and functional confounds of inter-piriform coupling: Panel A.** shows the x (top), y (middle) and z (bottom) MNI coordinates of the left and right piriform segmentations after they were registered to MNI space (using the T1w image) for the control, high and low IPC groups. Similarly **Panel B.** shows the piriform volumetry split by those same groups. **Panel C.** shows the temporal signal-to-noise ratio (tSNR) of the BOLD signal within the piriform ROI across the same groups. **Panel D.** shows the correlation between the mean BOLD signal between the two piriform ROIs in native space, as a direct proxy for IPC (Spearman's $r$ = 0.67, $p<0.001$). n.s. - not significant. *p<0.05, **p<0.01, ***p<0.001, ****p<0.0001. $p$-values are Bonferroni corrected.

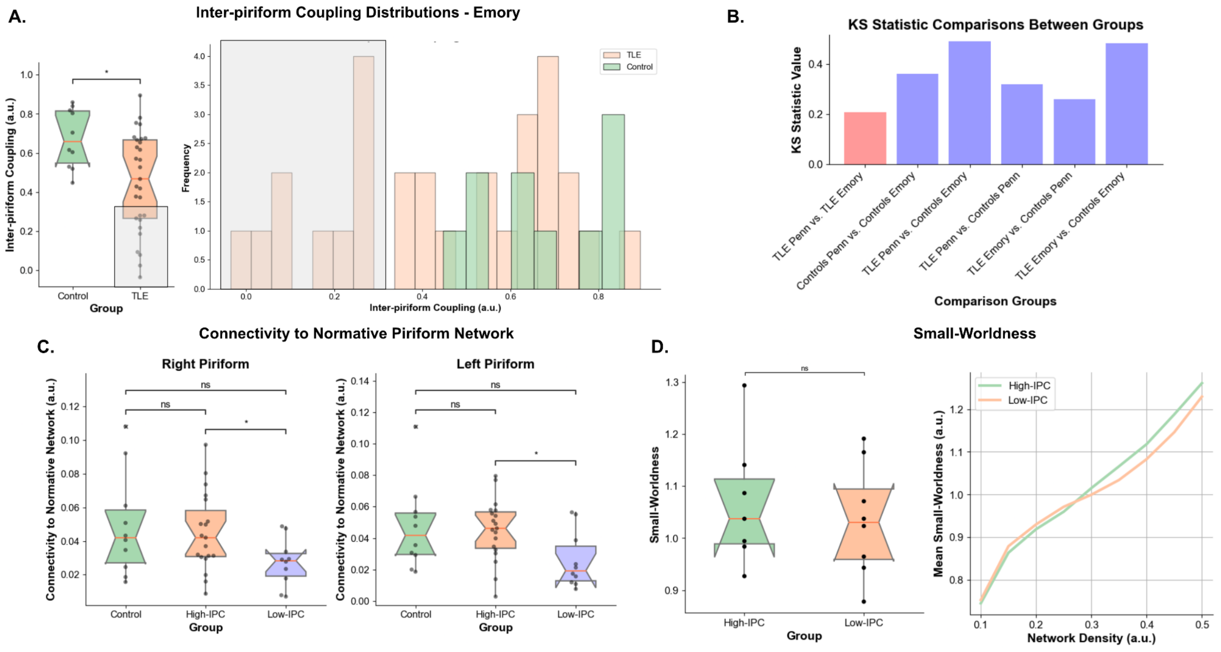


**Supplementary Figure 8 - Inter-piriform coupling in external validation dataset: Panel A.** shows the inter-piriform coupling for healthy control and TLE subjects in the Emory dataset. We also notice a bimodal distribution within the TLE group, with a clear separation between the Low-IPC and High-IPC subjects (gray rectangle). This separation happens at a value of 0.30 in this cohort. **Panel B.** shows the value of the Kolmogorov-Smirnov statistic after doing a pairwise comparison of the distributions of our original TLE cohort (TLE Penn), the Emory TLE cohort (TLE Emory), the original control cohort (Controls Penn) and the Emory control cohort (Controls Emory). **Panel C.** shows the connectivity of the piriform to the normative piriform connectivity mask reported in the original manuscript. **Panel D.** shows the small-worldness estimated for the subset of Emory subjects that had rs-fMRI acquisitions longer than 5 minutes (a subset of the included Emory subjects had rs-fMRI acquisitions lower than 5 minutes, which is known to cause instability in rs-fMRI network metrics).

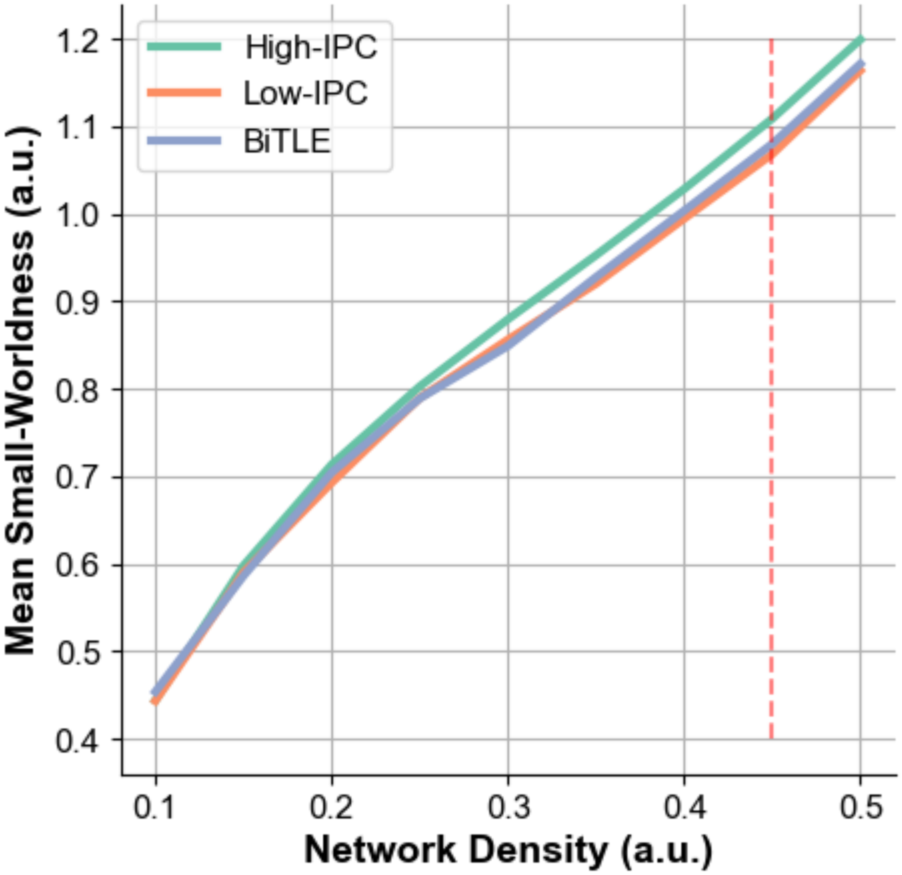


**Supplementary Figure 9 - Small-worldness as a function of network density:** We show the mean small-worldness across subjects in each group for a range of network densities between 0.1 and 0.5. The red line is the threshold density of 0.45 which is the value used in the main results of the manuscript.
